# Integrative genomic analyses in adipocytes implicate DNA methylation in human obesity and diabetes

**DOI:** 10.1101/2021.12.20.21266893

**Authors:** L. McAllan, D. Baranasic, S. Villicaña, W. Zhang, B. Lehne, M. Adamo, A. Jenkinson, M. Elkalaawy, B. Mohammadi, M. Hashemi, Y. Yang, L. Zudina, V. Lagou, S. Tan, J. Castillo-Fernandez, R. Soong, P. Elliott, J. Scott, M. Loh, I. Prokopenko, B. Lenhard, R. L. Batterham, J. Bell, J. Chambers, J. Kooner, W. Scott

## Abstract

DNA methylation variations are prevalent in human obesity, but evidence of a causative role in disease pathogenesis is limited. In this study, we combine epigenome-wide association and integrative genomics to investigate the impact of subcutaneous and visceral adipocyte DNA methylation variations in extreme human obesity. We identify extensive DNA methylation changes that are robustly associated with extreme obesity in combined discovery and replication analyses (N=190 samples, 691 loci in subcutaneous and 173 loci in visceral adipocytes, P<1×10-7). Using functional interaction maps and methylation-expression association testing in human adipocytes, we connect extreme obesity-associated methylation variations to transcriptomic changes at >500 target genes. We find that disease-associated methylation variations localise to active genomic regions and transcription factor binding sites, at which DNA methylation influences transcription factor-target gene co-expression relationships. In Mendelian Randomisation analyses, we infer causal effects of DNA methylation on human obesity and obesity-induced metabolic disturbances, under genetic control, at 28 independent loci. Silencing of two target genes of causal DNA methylation variations, the *PRRC2A* and *LIMD2* genes, further reveals novel metabolic effects in adipocytes. Our results indicate DNA methylation is an important determinant of human obesity and its metabolic complications, and reveal genomic and molecular mechanisms through which altered DNA methylation may impact adipocyte cellular functions.

## INTRODUCTION

Obesity is a disease of excess adipose tissue that impairs health^1^. Worldwide there are more than 650 million people affected by obesity^2^. These individuals are at high risk of developing obesity-induced inflammatory and metabolic disturbances, and subsequent type 2 diabetes (T2D)^3–5^. Existing treatments for obesity and T2D have major limitations^6^, and new therapeutic targets derived from increased understanding of disease mechanisms are a global health priority.

DNA methylation, the first layer of epigenetic regulation, is causally implicated in human obesity and T2D through diverse aetiological pathways^7–9^. These pathways include in utero programming of disease risk, long-term effects of diet and lifestyle, mediation of causal genetic variations, age-related susceptibility and even inter/trans-generational inheritance^10–16^. However, previous efforts to identify robust causal associations between DNA methylation and these common harmful human conditions have been hindered by issues with tissue selection, cell specificity and assigning causation^17–21^. Obesity-associated DNA methylation changes in human blood are primarily a consequence rather than a cause of disease^22, 23^. Methylation variations in human metabolic tissues cannot be confidently linked to phenotype due to cellular and therefore epigenetic heterogeneity^18, 19^. Very few studies have investigated genome-wide DNA methylation in clinically relevant human cell-types due to the substantial challenges in collecting and then isolating cells from human tissues^24–26^.

Adipocytes are the major cell-type in adipose tissues. These specialist metabolic cells have important roles in local energy storage and expenditure, whole-body energy and glucose homeostasis, and obesity and T2D pathogenesis^27, 28^. Interestingly, adipocytes in distinct anatomical locations have variable DNA methylation and transcriptomic profiles, molecular functions and impacts on metabolic health, with visceral adipocytes considered more harmful than subcutaneous adipocytes^25, 29, 30^. Recent experimental studies demonstrate that manipulation of DNA methylation enzymes in adipocytes can induce or prevent obesity and T2D, through cellular effects on energy expenditure and insulin sensitivity^31, 32^. These proof-of-principle studies provide strong rationale for exploratory epigenomic studies in human adipocytes.

Here, we addressed key limitations of previous human epigenome-wide association studies (EWAS) in subcutaneous and visceral adipocytes with the aim of identifying novel epigenetic mechanisms underlying obesity or its adverse metabolic consequences. We used an integrated genomic strategy to: i. identify robust alterations in human adipocyte DNA methylation associated with extreme obesity; ii. predict the potential effector transcripts (cis-target genes) of these DNA methylation changes; and iii. infer mechanisms underlying these DNA methylation-gene expression relationships. At a subset of methylation sites and target genes, we used genetic association and adipocyte genetic manipulation to provide complementary evidence of causation. Our results highlight the importance of studying cell type-specific epigenomic variations and the power of extreme trait sampling. We provide mechanistic insights into the role of DNA methylation in human obesity and T2D, and deliver novel targets for detailed functional characterisation and potential clinical translation.

## RESULTS

### Genome-wide alterations in adipocyte DNA methylation in people with extreme obesity

To identify obesity-associated alterations in human adipocyte DNA methylation (5-methylcytosine, 5mC) we collected subcutaneous and visceral adipose tissue samples intraoperatively from people with extreme obesity and healthy controls, and isolated populations of adipocytes from these tissues. We then characterised genome-wide DNA methylation in 190 subcutaneous and visceral adipocyte samples from obese cases and controls in separate discovery and replication cohorts (Illumina 450K and EPIC Beadchips, **Supplementary Figure 1**). The mean difference in body mass index (BMI) between obese cases and controls from both discovery and replication cohorts was large (∼20kg/m2) (**Supplementary Table 1**; age (±3.5-yrs), sex and ethnicity matched).

In subcutaneous adipocytes, we discovered 4485 5mC sites associated with extreme obesity at a false discovery rate (FDR) of <1%^33, 34^. We then replicated the association of 5mC with obesity at 905 of these sites in an independent subcutaneous adipocyte sample (at FDR<1% in the replication sample, and at epigenome-wide significance P<1×10-7 in the combined discovery and replication samples, Fig. 1A). For subsequent genomic and functional analyses, we annotated the sentinel 5mC site at each replicated genomic locus (691 subcutaneous sentinels, lowest combined discovery and replication P value, ≥5-kb apart which is equivalent to reported CG island (CGI) widths, **Supplementary Table 2**). Subcutaneous adipocyte sentinels had a median 5.8% (range 1.1-17.9%) difference in methylation between obese cases and controls, and were systematically hypomethylated in obese cases (binomial sign test P<6.4×10-33, Fig. 1B, consistent with previous reports^35^). More sentinels had intermediate (0.2-0.8) than low (<0.2) or high (>0.8) methylation levels (Fig. 1B). 24 loci had ≥5 differentially methylated 5mC sites immediately flanking and within ±5-kb of the sentinel 5mC site (consistent association direction, P<0.05, Bonferroni adjusted), indicating extended regions of differential methylation (**Supplementary Table 2**). The largest differentially methylated regions were found at the *RUNX3* (19 sites, 1174-bp), *TBX5* (17 sites, 3187-bp) and *ISLR2* (14 sites, 4050-bp) loci. Overall, 4363 of the 4485 5mC sites identified at FDR<1% in the discovery sample showed directional consistency for association with extreme obesity in the replication sample (binomial sign test P<1×10-300), suggesting our findings represent only the strongest signals among a larger number of obesity-associated DNA methylation changes in subcutaneous adipocytes.

**Figure 1:**
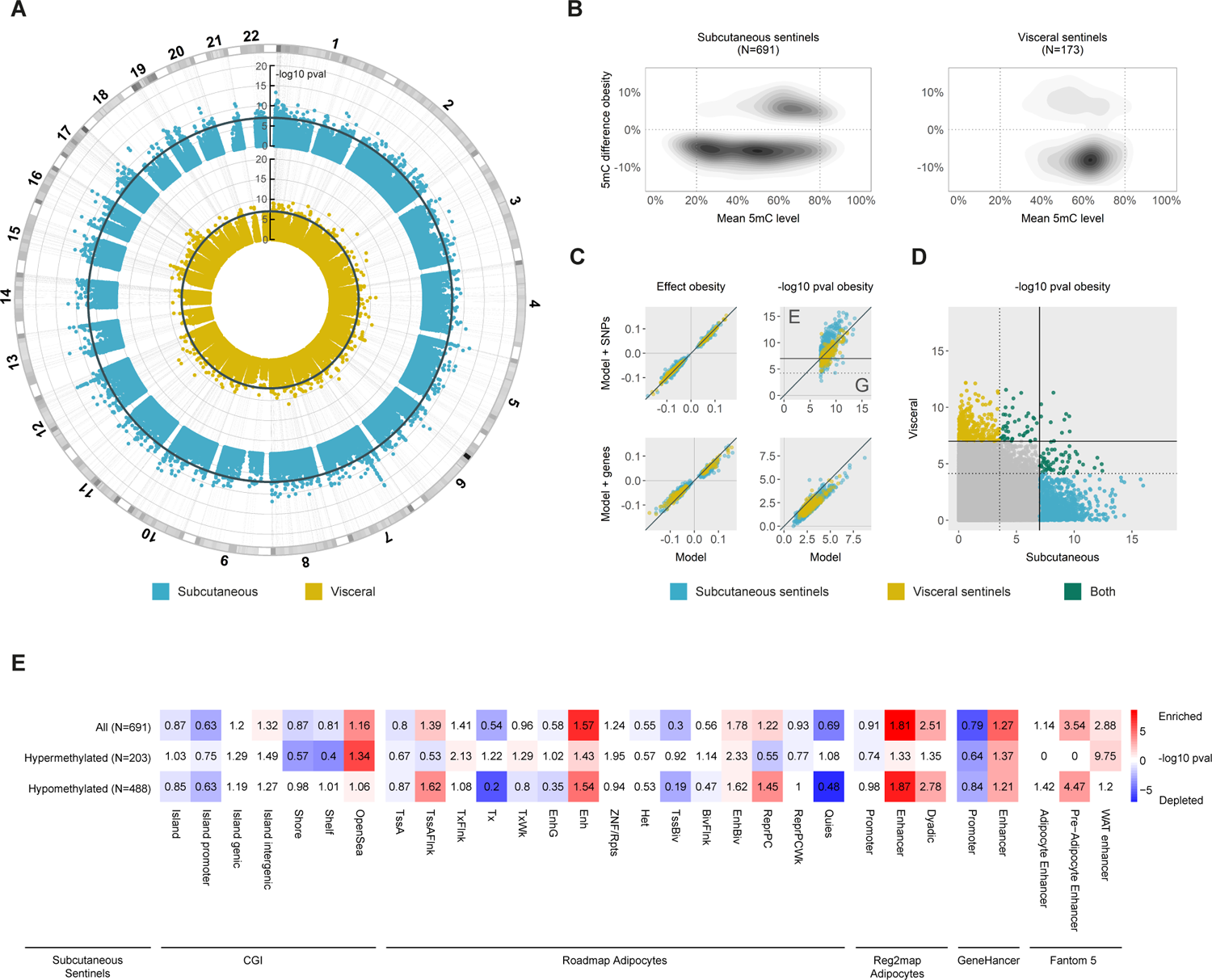
DNA methylation sites associated with extreme human obesity in subcutaneous and visceral adipocytes. **A.** Genome-wide associations between 5mC and extreme obesity in subcutaneous and visceral adipocytes (N=401,595 sites); −log10 pvalue in combined discovery and replication samples ordered by autosomal chromosome; threshold line epigenome-wide significance (EWS, P<1×10-7). **B.** 5mC differences between obese cases and controls relative to mean 5mC levels at N=691 subcutaneous and N=173 visceral sentinel methylation sites (%-methylation). **C.** Comparisons of association models without and with adjustment for potential confounding variables to evaluate the effects of genetic variations and potential contaminating cell genes on 5mC-obesity relationships. Top panels: adjustment for cis-SNPs associated with each sentinel 5mC site (FDR<0.01); effect size (beta) and −log10 pvalue in the combined discovery and replication cohorts; solid threshold EWS; dashed threshold Bonferroni corrected pvalue (0.05/N sentinels); G genetic effects; E non-genetic effects. Bottom panels: adjustment for principal components (PC1-5) derived from expression of 12 potential contaminating cell genes; effect size (beta) and −log10 pvalue in the replication cohort. **D.** Cross tissue effects. Methylation-obesity association pvalue in subcutaneous and visceral adipocytes (combined discovery and replication samples); solid threshold EWS; dashed threshold Bonferroni corrected pvalue (0.05/N sentinels). **E.** Genomic annotation of obesity-associated DNA methylation sites. Numbered by fold change (observed compared to mean expected) and coloured by enrichment (red) or depletion (blue) −log10 pvalue (Fishers Exact Test).

In visceral adipocytes, we identified 445 5mC sites associated with obesity at FDR<1%, markedly fewer sites than in subcutaneous adipocytes. 220 of these 5mC sites replicated in an independent visceral adipocyte sample (replication FDR<1%, combined discovery and replication P<1×10-7, Fig. 1A). The 173 sentinel sites (visceral sentinels, lowest P-value, ≥5-kb apart) had median methylation difference of 7.9% (range 2.9%-21.5%) between obese cases and controls (Fig. 1B, **Supplementary Table 3**). Visceral adipocyte sentinels were also preferentially hypomethylated in obesity (P=3.8×10-7), and most sentinels had intermediate (0.2-0.8) methylation levels (Fig. 1B). 2 loci showed extended regions of differential methylation (≥5 significantly differentially methylated 5mC sites within ±5-kb of the sentinel site, **Supplementary Table 3**); at *NFIA* (5 sites, 2081-bp) and near *ATG5* (5 sites, 831-bp). Again, overall directions of effect in visceral adipocytes were highly concordant between the discovery and replication samples (440 of 445 5mC sites, P<2×10-123).

Disease-associated alterations in DNA methylation are frequently the result of underlying differences in genetic polymorphisms or cellular (and therefore epigenetic) heterogeneity between individuals^18, 19^. In sensitivity analyses, we found that the majority of sentinels remained associated with obesity after correction for genetic effects (Fig. 1C**, Supplementary Figure 2**). Consistent with this, <1% of sentinels showed methylation distributions fitting with underlying SNP effects^36^, indicating that the identified methylation differences are predominantly environmentally driven. Similarly, we found that potential cellular heterogeneity (arising from impurity) and other unmeasured confounding exposures did not systematically alter our findings (Fig. 1C**, Supplementary Figures 3 and 4**).

As adipose tissues from different depots have varying impacts on metabolic health^29^, we examined whether 5mC changes associated with obesity were specific to subcutaneous or visceral adipocytes. Only 23 subcutaneous adipocyte sentinels were robustly associated with obesity in visceral adipocytes, while a further 11 visceral sentinels replicated in subcutaneous adipocytes (consistent direction and P<0.05, Bonferroni corrected for the number of sentinels, Fig. 1D, **Supplementary Table 4**). Similarly, we found only weak concordance when we compared overall directions of effect between subcutaneous and visceral adipocyte sentinels (374 of 671, P=0.003 subcutaneous in visceral, and 105 of 173 P=0.006 visceral in subcutaneous).

Together, these genome-wide discovery and replication analyses quantify extensive alterations in DNA methylation in adipocytes from people with extreme obesity. Surprisingly, the majority of extreme obesity-associated 5mC changes are adipose depot specific, raising the possibility of tissue intrinsic biological origins and functions.

### Enrichment of extreme obesity-associated 5mC sites in active genomic regions

The genomic location of DNA methylation influences its gene regulatory potential and its mechanism of action^9, 37, 38^. To identify 5mC sites with active gene regulatory potential and evaluate underlying mechanisms, we mapped obesity-associated subcutaneous and visceral adipocyte sentinels to the human reference genome, human CpG island (CGI) annotations, and human adipose/adipocyte functional genomic maps.

Differentially methylated subcutaneous adipocyte sentinels were significantly enriched in adipose tissue and adipocyte enhancers (Roadmap chromatin states, Reg2Map, Fantom5^39–41^) and enhancers predicted from multifaceted datasets (Genehancer^42^, Fig. 1E**, Supplementary Figure 5**). Enrichment in enhancers was explained by both hypo-(lower methylation in obese adipocytes) and hyper-(higher methylation in obese adipocytes) methylated subcutaneous sentinels. Hypo-methylated sentinels were also (weakly) enriched in regions flanking active transcription start sites (TSS), bivalent enhancers and repressed polycomb, whereas hyper-methylated sentinels were generally underrepresented in these regions. In contrast, hyper-methylated sentinels were enriched and hypo-methylated sentinels were diminished in actively transcribed genic regions. In addition, subcutaneous sentinels were under-represented at promoter CGIs but not at intra-or inter-genic CGIs (Fig. 1E**, Supplementary Figure 5**). Differentially methylated visceral adipocyte sentinels showed similar trends in CGIs and multifaceted enhancer datasets but were not significantly enriched in these or in adipose tissue/adipocyte chromatin states (which notably are derived from the subcutaneous adipose depot, **Supplementary Figure 5**). Our findings are consistent with previous studies localising variably methylated regions to cis-regulatory regions involved in transcriptional control and phenotypic variation^43–46^, rather than CG dense DNA sequences in the promoters of core housekeeping genes.

We also examined whether the genomic regions flanking our 5mC sentinels contained DNA sequence variants associated with human obesity phenotypes in GWAS. Although we found no evidence of enrichment, 11 loci contained genetic variants associated with obesity (BMI^47^), 8 central adiposity (waist-hip-ratio adjusted for BMI, WHR^48^) and 4 type-2 diabetes (T2D^49^) at genome-wide significance (P<5×10-8, **Supplementary Table 5**); these loci included the BMI and adiposity locus *NRXN3*^50, 51^, the WHR locus *TBX15*^52, 53^ and the T2D locus *TCF7L2*^54^.

### Predicted target genes of extreme obesity-associated 5mC sites

To identify genes that might be responsible for the effects of DNA methylation on phenotype (target effector genes), we carried out RNA sequencing in obese and lean subcutaneous and visceral adipocytes with paired DNA methylation results (replication cohort, N=89 samples, median 49M assigned reads). We split our 5mC sentinels into those in gene promoters, 5/3’UTRs and exons (N=389 subcutaneous and N=92 visceral sentinels) with unambiguous target genes, and those in intronic and distal intergenic regions (N=302 subcutaneous and N=81 visceral sentinels) without defined target genes.

At promoters, 5/3’UTRs and exons, we examined the relationship between change in methylation at the 5mC sentinel site and change in expression of the overlapping/directly flanking cis-genes (mixed-effects model linear regression, combined subcutaneous and visceral adipocyte samples). Methylation at 121 subcutaneous adipocyte sentinels was robustly associated with expression of 126 unique cis-genes at FDR<0.01 (P range 9.1×10-3 to 1.9×10-24, Fig. 2A and 2D, **Supplementary Table 6**). In addition, methylation at 29 visceral adipocyte sentinels was associated with expression of 32 unique cis-genes at FDR<0.01 (P range 4.6×10-3 to 1.6×10-12, Fig. 2D, **Supplementary Table 7**). Methylation changes at promoters, 5/3’UTRs and exons had predominantly negative effects on gene transcription (Binomial test P=0.004, Fig. 2E). Subcutaneous sentinels associated with gene transcriptional changes were enriched in the genomic regions flanking active TSS rather than at active TSS (2.4-fold, Fisher’s exact P=0.0005 in adipocytes and 2.4-fold, P=7.4×10-5 in adipose, Fig. 2E**, Supplementary Figure 6**).

**Figure 2:**
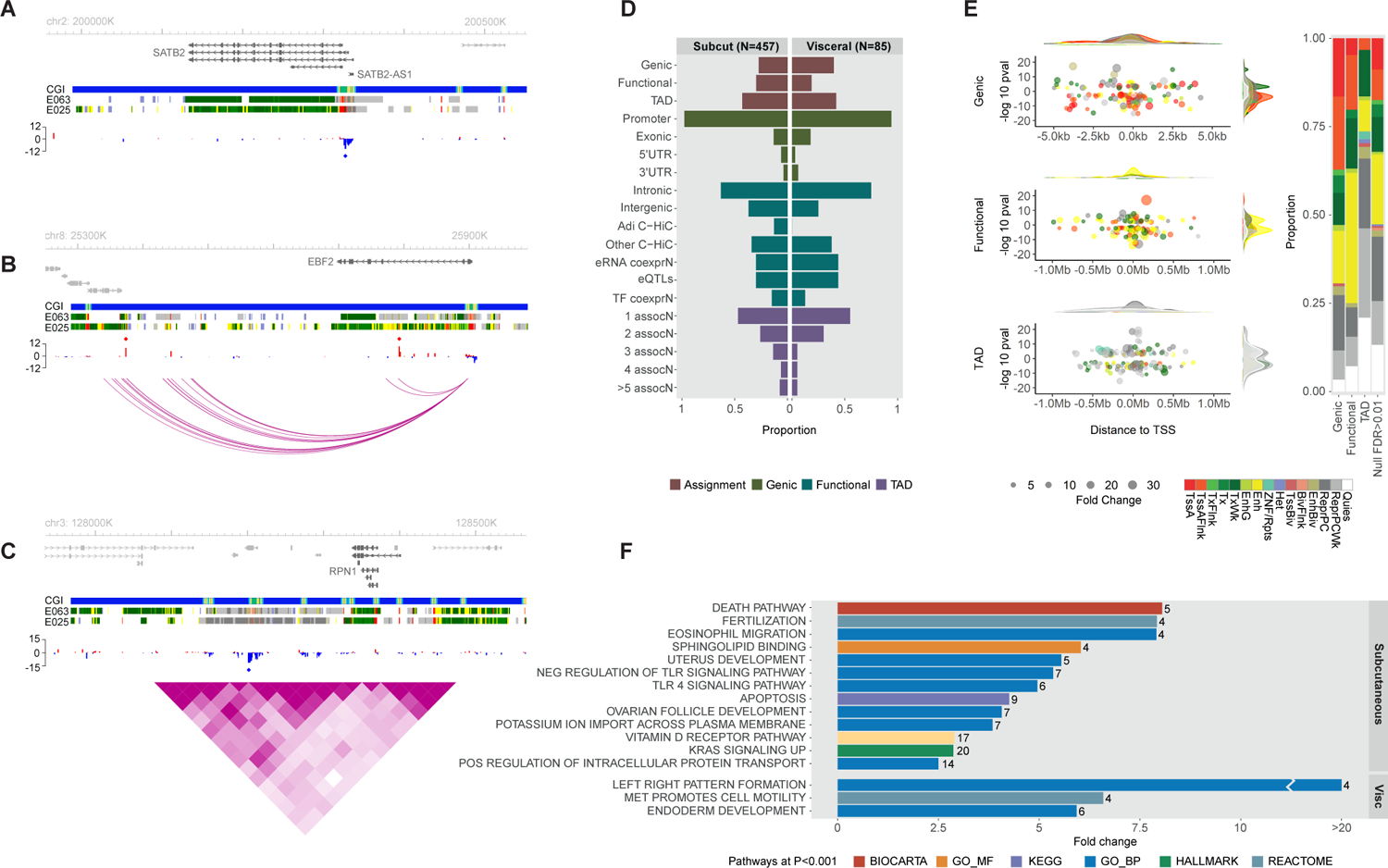
DNA methylation-target gene associations in human adipocytes. **A-C.** Locus plots of sentinel 5mC sites (diamond) and their predicted target effector genes (dark grey). **A.** Methylation at cg01558212 in the promoter of *SATB2* was associated with *SATB2* and *SATB2-AS1* gene transcription (subcutaneous). **B.** Methylation at two sites, cg11307296 and cg13390388 within distinct functional loops in human adipocyte promoter capture HiC connectivity maps, was associated with transcription of the adipocyte browning/beigeing gene *EBF2* (subcutaneous). **C.** Methylation at cg03779326 was associated with transcription of *RPN1* but not other putative target genes within a shared human adipocyte TAD (visceral). Presented as %-difference in methylation between obese cases and controls and annotated by UCSC CpG island (CGI) and Roadmap adipose (E063) and adipocyte (E025) chromatin states. **D.** Frequency of sentinel methylation-expression associations at FDR<0.01 according to their target gene assignment method. Genic: sentinel in a promoter, 5/3’UTR or exon. Functional: intronic/intergenic sentinel sharing a functional interaction with a distal target gene. TAD: intronic/intergenic sentinel and distal target gene(s) with a shared human adipocyte topologically associated domain. Adi C-HiC: human adipocyte promoter capture HiC interaction. Other C-HiC: promoter capture HiC interaction in another human tissue. eRNA coexprN: co-expression of distal eRNA and proximal promoter RNA. eQTLs: Association of distal SNP with proximal promoter expression. TF coexprN: TF binding site in distal site (ChIP-seq) and TF-target gene co-expression. 1 to >5 assocN: Number of sentinel-target gene associations in a shared TAD. **E.** Subcutaneous sentinel-target gene associations at FDR<0.01 grouped by target gene annotation method, and coloured by adipocyte chromatin state (Roadmap E025). Left panel: distance to TSS and −log10 pvalue according to direction of effect, and sentinel density distribution. Right panel: frequencies of observed associations compared to the null background (sentinel-gene associations at FDR>0.01). Fold change: log2 fold change in gene expression for each unit change in DNA methylation. **F.** Enriched pathways and genesets at P<0.001 (Empirical) based on the nearest cis−gene to each 5mC sentinel in subcutaneous and visceral adipocytes. Bar represents fold change of observed compared to mean expected frequency, and number is the observed gene counts, in each pathway/geneset. All methylation-expression analyses were carried out in combined adipocyte samples.

At intergenic and intronic sites, we used human adipocyte chromosomal interaction maps (promoter capture Hi-C^55^) and enhancer-promoter inference datasets (GeneHancer^42^) to functionally assign 5mC sentinels to specific distal target genes, then tested sentinel 5mC sites for association with target gene expression. 84 subcutaneous and 14 visceral sentinels were associated with 135 and 16 unique distal target genes respectively at FDR<0.01 (P range 1.1×10-2 to 8.6×10-18, Fig. 2B and 2D**, Supplementary Tables 8 and 9**). For intergenic and intronic sentinels not assigned to target genes through direct functional interactions (N=161 in subcutaneous and N=56 in visceral) we localised the 5mC site to a human adipocyte topologically associated gene regulatory domain (TAD^56^), and defined all genes within the domain as potential targets (median 7, range 1-46 target genes per sentinel). 91 subcutaneous sentinels were associated with 177 cis-genes and 21 visceral sentinels with 34 cis-genes in the same TAD at FDR<0.01 (P range 2.7×10-3 to 3.5×10-20, Fig. 2C and 2D**, Supplementary Tables 10 and 11**). Most associations occurred in unique tads, though 8 TADs had ≥5 sentinel methylation-target gene associations (Fig. 2D). As expected, sentinels associated with functionally assigned target genes were enriched in enhancers (subcutaneous 1.8-fold P=8.4×10-4 in adipocytes and 1.5-fold P=0.03 in adipose; visceral 3.1-fold P=0.05 in adipocytes and 3.3-fold P=0.01 in adipose, Fig. 2E and **Supplementary Figure 6**). By contrast, sentinels associated with genes in a shared TAD were enriched in polycomb repressed regions (subcutaneous 1.5-fold P=0.007 in adipocytes, 1.8-fold P=0.0002 in adipose, Fig. 2E), suggesting that genomic context-specific regulatory mechanisms underlie the observed methylation-expression associations.

Many of the genes associated with altered DNA methylation have important roles in insulin signalling/sensitivity, adipogenesis, fatty acid metabolism and adipocytokine signalling (**Supplementary Tables 6 to 11**). As examples, the *IRS2*, *ADIPOR2*, *PLIN5*, *FABP3, SOCS3, RBP4* and *AKT3* genes in subcutaneous adipocytes^57–63^, and the *KLF6*, *TULP3, DOC2B*, *ACOT11*, *PRKD2*, *SLC22A3* and *KIF5C* genes in visceral adipocytes^64–70^. Interestingly, several methylation-expression associations involved genes that control neurite formation, axonal guidance and synaptic plasticity (*NRN1*, *SEMA6B*, *SEMA4B* and *NRXN3*^71–73^), raising the possibility of epigenetic effects on neural inputs to expanding adipose tissue. Subcutaneous sentinels were associated with important genes involved in browning/beigeing of white adipocytes including the master regulator transcription factors *PRDM16* and *EBF2*^74, 75^, and the signalling molecules *FGF9* and *IL10RA*^76, 77^, though directions of effect on browning were inconsistent. We also discovered a novel association between expression of a cluster of micro-RNAs implicated in adipogenesis, MIR23A, MIR24-2 and MIR27A^78–83^, and methylation at its shared promoter in subcutaneous adipocytes. Nonetheless, most potential effector genes of obesity-associated 5mC changes have unknown functions in adipocytes.

### Functional annotation of target genes linked to extreme obesity-associated 5mC changes

We used gene set enrichment analyses to systematically evaluate the molecular and functional significance of cis-genes linked to our 5mC sentinels either by genomic location (nearest cis-gene) or by association (methylation-expression FDR<0.01). The nearest cis-genes to subcutaneous sentinel 5mC sites were enriched in growth and development, inflammatory, metabolic and apoptosis pathways (FDR<0.01 Empirical, Fig. 2F, **Supplementary Table 12**). Notable pathways included *TLR4* signalling, a key trigger of the obesity-induced inflammatory response^84^, sphingolipid binding which is implicated in cell stress and metabolic dysfunction^85^, and Vitamin D metabolism which is linked to adipogenesis, lipid storage and adipocytokine production^86^. The genes nearest to visceral sentinels were over-represented in endoderm development, body pattern formation and cell motility pathways (FDR<0.01 Empirical, Fig. 2F, **Supplementary Table 12**). Cis-genes associated with change in methylation at subcutaneous sentinels were enriched in a cluster of intersecting gene sets involved in transcriptional control and cell/tissue development (FDR<0.01 Hypergeometric, gProfiler^87^). Relevant sub-clusters included cell differentiation and muscle development gene sets (**Supplementary Figure 7 and Table 13**). Similarly, cis-genes associated with visceral sentinels were enriched in intersecting embryonic and tissue development gene sets, and in transmembrane drug transporter genes (FDR<0.01 Hypergeometric, gProfiler, **Supplementary Figure 7 and Table 13**). Taken together, these pathway analyses link obesity-associated 5mC sites in subcutaneous and visceral adipocytes to genes involved in cell lineage and fate determination, tissue development and remodelling, inflammation and metabolic function/dysfunction.

### Evidence of mechanistic interactions between transcription factors and extreme obesity-associated 5mC sites

Recent studies suggest that DNA methylation in enhancers and other active cis-regulatory regions may systematically regulate gene transcription by altering the binding of methylation-sensitive transcription factors (TFs^88–90^). Alternatively, TF binding can alter the methylation status of flanking DNA in these regions^89, 90^. To investigate putative mechanistic interactions between obesity-associated 5mC changes and TFs, we mapped human TF binding motifs within ±150-bp of each sentinel 5mC site using the Homer database^91^. We then identified the TFs with the strongest potential to bind to each motif, and examined the relationship between expression of these TFs, change in sentinel methylation and change in sentinel target gene transcription in adipocytes.

The genomic regions flanking the 671 subcutaneous sentinels were enriched for 7 distinct TF binding motifs (P range 1×10-9 to 1×10-13); 5 motifs at hypo-(lower in obesity) and 2 motifs at hyper-methylated (higher in obesity) sentinels (median 28 range 16-86 sentinels per motif, Fig. 3A, **Supplementary Table 14**). Motif 1 was preferentially located at tissue-specific enhancers, Motif 4 at regions flanking active TSS, and Motif 5 at both enhancers and active TSS flanking regions (Fig. 3B**, Supplementary Figure 8**). Overall, 49 TFs with potential to bind at these 7 motifs were expressed in human subcutaneous adipocytes (median 8 range 4-11 TFs per motif, **Supplementary Table 15**). In visceral adipocytes, we identified several putative motifs at obesity-associated 5mC sites (**Supplementary Table 16**) but none reached the stringent significance threshold required for robust de novo motif discovery.

**Figure 3:**
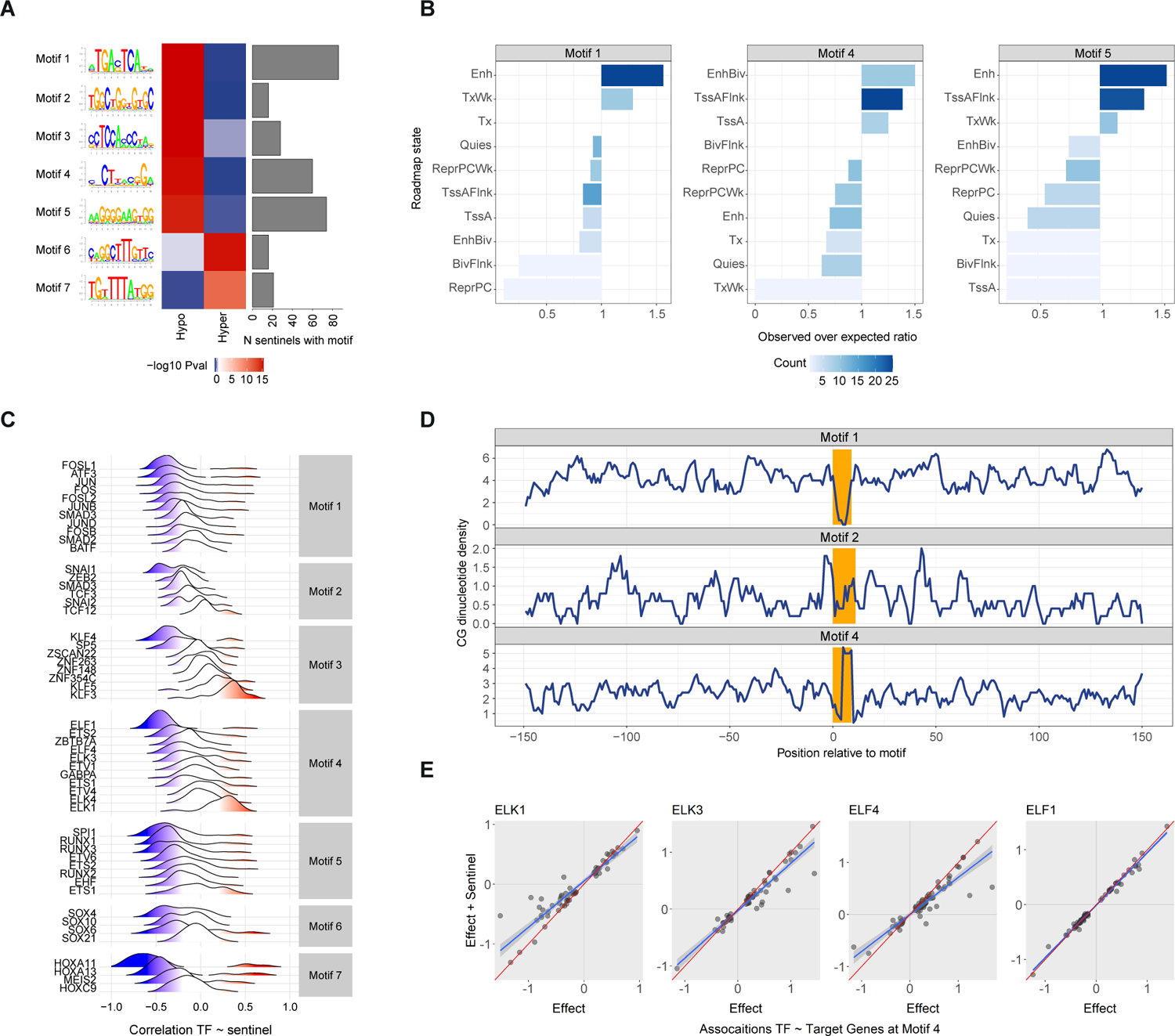
Interactions between DNA methylation and transcription factors in human subcutaneous adipocytes. **A.** Enrichment of extreme-obesity associated DNA methylation sentinels in 7 transcription factor binding motifs (subcutaneous sentinels). Left panel: the predicted DNA sequence corresponding to each enriched motif, based on observed nucleotide frequencies (Homer). Motifs 2 and 4 both contained CG sites within their predicted DNA binding sequence. Centre panel: heatmap of −log10 pvalue for enrichment of: i. hypo-(lower in obesity); and ii. hyper-methylated subcutaneous sentinels (relative to permuted background, hypergeometric distribution). Right panel: bar plot of the number of subcutaneous sentinels mapping to each motif. **B.** Human adipocyte Roadmap chromatin state annotation of Motifs 1, 4 and 5; bar plots of observed over expected ratio in selected roadmap states, coloured by observed counts (number of sentinel-motif pairs). Motif 1 was over-represented in enhancers, Motif 4 in active TSS, and Motif 5 in enhancers and active TSS. **C.** Density/ridge plots of pairwise correlation between TF expression and methylation level at each of its corresponding sentinels in subcutaneous adipocytes, split by Motif and ranked by mean correlation. **D.** Distribution of genomic CG sites in the ±150-bp regions flanking Motifs 1, 2 and 4, centred on the motif (coloured in orange). Genomic CG sites were strongly enriched at Motif 4 (peak), and strongly depleted at Motif 1 (trough), relative to the flanking DNA sequences. A peak of genomic CGs was also observed immediately upstream of Motif 2 though other genomic CG peaks were present in its flanking DNA sequences. **E.** Relationships between expression of 4 TFs predicted to bind at Motif 4 (*ELK1*, *ELK3*, *ELF1*, *ELF4*) and expression of the assigned target genes of each methylation sentinel corresponding to Motif 4. Presented as association beta without (Effect) and with (Effect + Sentinel) adjustment for sentinel DNA methylation level (combined adipocyte samples). Adjustment for sentinel DNA methylation levels systematically influenced the associations between the *ELK1*, *ELK3* and *ELF4* TFs and their target genes, but not the *ELF1* TF.

Consistent with evidence that TF binding activity can alter DNA methylation levels, we found correlation between TF expression and sentinel methylation levels at each motif (Fig. 3C). The strongest correlations involved the *HOXA11* and *HOXA13* TFs (Motif 7, Fig. 3C), with weaker correlations between the *ATF3, FOSL1* and *JUN* (Motif 1), *SNAI1* (Motif 2), *KLF3*, *KLF4* and *KLF5* (Motif 3), *ELF1* and *ELK1* (Motif 4), *RUNX1*, *RUNX3* and *SPI1* (Motif 5), and *SOX4*, *SOX6* and *SOX10* (Motif 6) TFs and subsets of their sentinels (Fig. 3C). Motifs 2 and 4 contained CG sites within their binding sites, raising the possibility that the presence of DNA methylation might directly alter TF binding affinity at loci linked to these motifs (Fig. 3A, **Supplementary Table 14**). To evaluate this in more detail, we examined whether these motifs were enriched for differentially methylated sites (sentinel and flanking sites) associated with obesity. No enrichment was observed but array coverage of potential CG sites was sparse (Median 22% IQR 14-33% of genomic CG sites within ±150bp of a Motif, **Supplementary Figure 8**). As DNA methylation levels at adjacent CG sites are correlated^92^, we examined motifs for enrichment of genomic CG sites not covered in our dataset at which 5mC could impact TF-DNA binding. Motif 4 was strongly enriched for genomic CG sites relative to the flanking DNA sequences (±150-bp, Fig. 3D). A cluster of genomic CGs was also present immediately upstream of Motif 2, though other genomic CG clusters were observed in the DNA sequences flanking Motif 2. We therefore examined whether sentinel methylation levels at Motifs 2 and 4 influenced co-expression between TFs and their predicted target genes (the predicted target genes of each sentinel site corresponding to that TF-Motif pair). At both motifs, levels of methylation impacted TF-target gene relationships, suggesting potential mediation of transcriptional regulation by methylation (Fig. 3E**, Supplementary Figure 8**). The largest effects of methylation on TF-target gene relationships were observed at the *ELK1*, *ELK3* and *ELF4* TFs (Fig. 3E), of which *ELK1* and *ELF4* have been shown to be methylation-sensitive *in vitro*^93^. Importantly, the TFs implicated in these reciprocal relationships with DNA methylation are widely involved in adipocyte biology, adiposity and metabolic dysfunction (**Supplementary Table 15**^53, 64, 94–103^).

### Genetic association analyses to infer disease causation

To distinguish individual 5mC sites with potential causal effects on obesity or obesity-induced metabolic disturbances, we carried out two sample Mendelian Randomisation analyses (MR). We used whole adipose tissue (N=588 samples, Twins UK^104–107^) rather than adipocytes to increase power to identify independent cis-SNPs associated with each sentinel 5mC site (within +/500-kb and pairwise r2<0.01) and selected a significance threshold of 0.05 (Bonferroni corrected) to reduce weak instrument bias. We then used the identified cis-SNPs as instrumental variables (IV) to infer causal effects of DNA methylation on obesity, central adiposity, T2D and glycaemic traits linked to T2D among large-scale human GWAS (Fig 4A^47–49, 108^).

**Figure 4:**
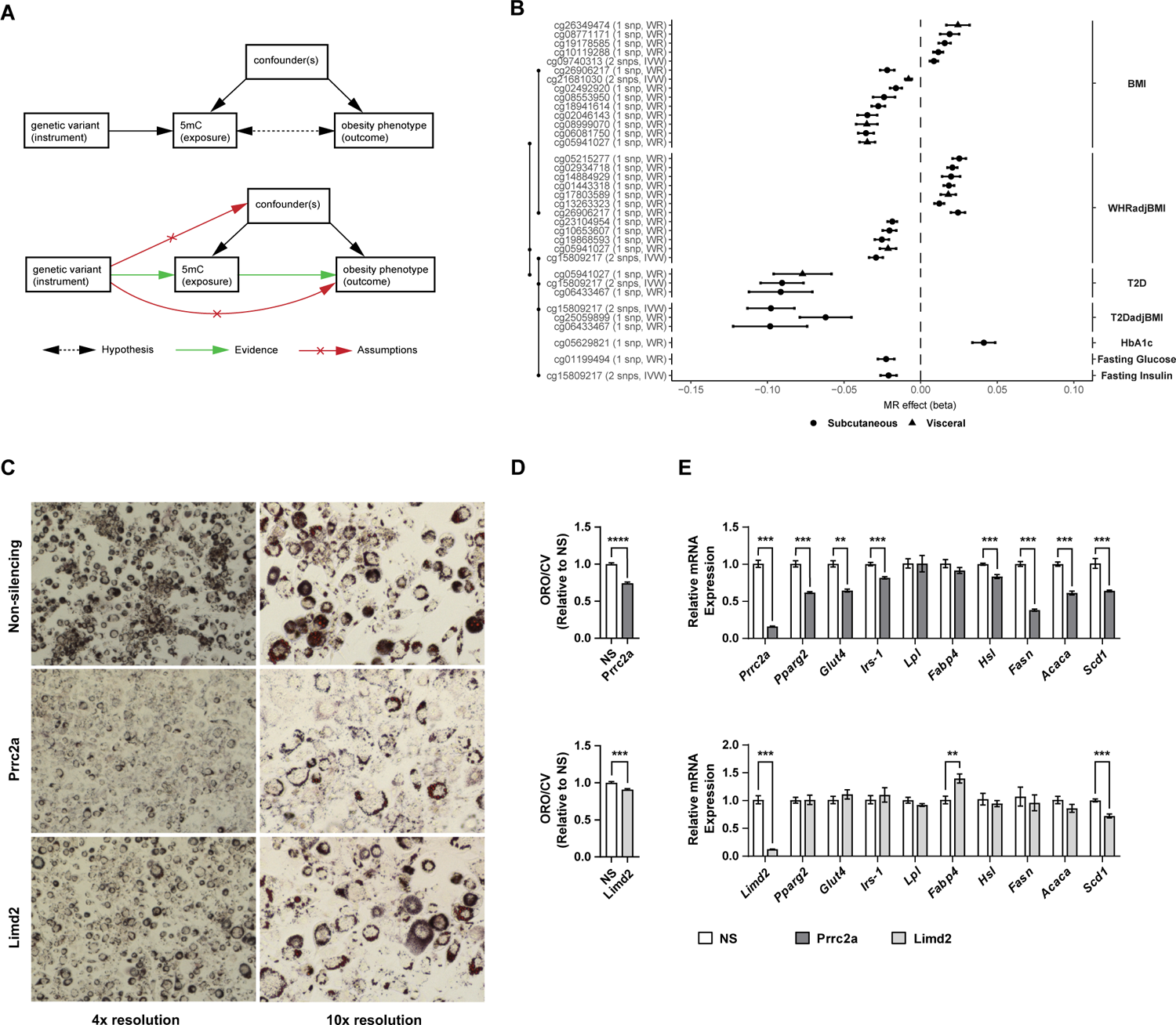
Causal inference and adipocyte functional analyses. **A.** Mendelian Randomisation analysis using genetic variants as instrumental variables to evaluate cause-consequence relationships between DNA methylation sentinels and human obesity phenotypes. Required evidence: robust association between: i. the instrumental genetic variant and the methylation exposure; and ii. the instrumental genetic variant and the outcome phenotype. Assumptions: i. the instrument only influences the outcome through the exposure not through any other pathway; and ii. the instrument is not associated with confounders. **B.** Forest plots of the effect sizes of subcutaneous and visceral adipocyte sentinels causally associated with human obesity phenotypes through two sample MR in adipocytes (FDR<0.01 in both MR causal and Steiger directionality tests). Centre values mark effect size estimates and error bars show the 95% confidence intervals. MR causal tests: Wald Ratio for single SNP IV (WR); Inverse Variance Weighted for >1 SNP IV (IVW). BMI: body mass index as a measure of obesity (GIANT, N≤795,640). WHRadjBMI: Waist-hip ratio adjusted for BMI as a measure of central adiposity (GIANT N≤694,649). T2D and T2D adjusted for BMI as measures of T2D risk (DIAGRAM, N≤231,422). Fasting glucose and insulin (MAGIC, N≤138,589) and HbA1c (MAGIC, N≤159,940) as measures of glycaemic traits linked to T2D. **C-E.** Adipocyte functional studies. **C** Oil Red O (ORO, red/brown) lipid staining at 4x and 10x magnification in day 6 differentiated 3T3-L1 adipocytes that were reverse transfected with non-silencing (NS), *Prrc2*a or *Limd2* siRNA at day 2 of differentiation. **D.** Equivalent spectrophotometric measurements of eluted ORO from siRNA treated 3T3-L1 adipocytes, normalised for cell number using measurements of crystal violet (CV, day 6, N=4). **E.** Expression of adipogenesis, insulin signalling and lipid metabolism genes at day 6 of differentiation in 3T3-L1 adipocytes transfected with siRNA against *Prrc2a*, *Limd2* or NS control at day 2 of differentiation (N=6). Real-time qPCR values were normalised to housekeeping genes (*Nono*, *Ywhaz*). ORO/CV and expression values are presented as mean ± SEM relative to NS control. **P<0.01, ***P<0.001, ****P<0.0001 versus NS control (Student’s t-test).

Only 191 subcutaneous and 34 visceral sentinels had significantly associated cis-SNPs, meaning we were unable to assess causation at a large fraction of sites. Nevertheless, we found genetic evidence to support causal effects of 5mC on obesity (measured using BMI) at 10 loci in subcutaneous adipocytes and 4 loci in visceral adipocytes (MR single instrument Wald Ratio or multiple instrument IVW random effects FDR<0.01, and Steiger directionality test FDR<0.01, Fig. 4B, **Supplementary Table 17**^109, 110^). We also identified potential causal effects of DNA methylation on central adiposity at 12 loci (10 subcutaneous, 2 visceral, measured using WHR), T2D at 4 loci (3 subcutaneous, 1 visceral), and fasting glucose, insulin and HbA1c at 3 loci (all subcutaneous, Fig. 4B, **Supplementary Table 17**). Methylation at 3 loci was causally linked to multiple obesity phenotypes: cg26906217 (BMI and WHR; near the PIK3C2A gene); cg15809217 (WHR, T2D and FI; associated with *PRRC2A* expression in subcutaneous adipocytes); and cg05941027 (BMI, WHR and T2D; associated with *LIMD2* expression in visceral adipocytes). Another notable locus was the 5mC site cg21681030, causally associated with BMI through MR, and respectively associated with expression of *RHOQ*, a glucose receptor trafficking gene^111, 112^, in our human adipocyte samples.

As our MR analyses were predominantly based on single SNPs as IVs, we repeated them using a larger number of correlated cis-SNPs (pairwise r2<0.8, SNP-methylation P<0.05, Bonferroni corrected) as IVs to evaluate for horizontal pleiotropy (Fig. 4B). 18 loci had at least 3 correlated IV SNPs enabling sensitivity testing at these loci. 15 loci replicated using MR IVW regression and 4 replicated using the less powerful MR Egger regression (P<0.05, **Supplementary Table 17**). Multiple loci showed evidence of heterogeneity between IVs, but only 4 loci violated the MR assumption of no horizontal pleiotropy (MR Egger intersect P<0.05, **Supplementary Table 17**). We then evaluated the relevancy of our MR findings to adipocytes by replication testing the SNP-5mC associations identified in whole adipose tissue in subcutaneous and visceral adipocytes. Despite the small sample size, 8 of 31 cis-SNPs replicated in adipocytes at P<0.05 (Binomial Sign Test P=0.0001) and 24 of 31 cis-SNPs had consistent directions of effect (Binomial Sign Test P=0.003, **Supplementary Table 18**). Thus, through genetic association, we infer causal effects of adipocyte DNA methylation on obesity or obesity-induced metabolic disturbances at up to 28 independent genomic loci.

### Adipocyte functional studies

Finally, we selected two target genes of 5mC sites implicated in disease pathogenesis through MR, but without known metabolic functions, for functional screening in adipocytes. We focused on the *PRRC2A* gene which was linked to central adiposity, insulin resistance and T2D, and the *LIMD2* gene which was linked to obesity, central adiposity and T2D. We evaluated the effect of silencing each target gene on a cellular model of adipogenesis, because adipogenesis has an important role in adipose tissue expansion and insulin sensitivity. We found that knockdown of both genes significantly reduced lipid accumulation during adipocyte differentiation, with a more marked effect observed after *PRRC2A* than *LIMD2* knockdown (Fig. 4C and 4D**, Supplementary Figure 9**). As adipocyte differentiation models are prone to variability, we independently replicated our lipid accumulation studies and observed concordant results (**Supplementary Figure 9**). We also examined the effects of *PRRC2A* and *LIMD2* gene silencing on key adipogenesis, lipid metabolism and insulin signalling genes. *PRRC2A* knockdown led to a consistent reduction in expression of *PPARG*, the master regulator adipogenic transcription factor, and concordant reductions in lipid metabolism and insulin signalling genes that are regulated by *PPARG* (Fig. 4E**, Supplementary Figure 9**). In contrast, *LIMD2* gene silencing did not alter *PPARG* expression (Fig. 4E**, Supplementary Figure 9**). Instead, *LIMD2* silencing led to increased expression of the fat mobilising genes *FABP4* and *HSL,* and reduction in expression of the lipid synthesis gene *SCD1*, though findings were inconsistent across replicates (Fig. 4E**, Supplementary Figure 9**). Overall, our integrative genomic and adipocyte functional studies are consistent with lower methylation levels at cg15809217 in obese subcutaneous adipocytes promoting insulin resistance and T2D, through reduced *PRRC2A* expression and impaired adipogenesis. They also suggest that reduced methylation at cg05941027 in obese visceral adipocytes may increase *LIMD2* expression and thereby increase lipid storage. *PRRC2A* is recently reported as a reader of N6-methyladenosine (m6A), the most abundant internal modification to mRNA, offering a novel mechanism through which its metabolic effects may be mediated^113^.

## DISCUSSION

Epigenetic processes are tissue and cell specific which has made investigating their roles in complex human diseases a major challenge. In this study, we combine integrated functional genomics with extreme trait sampling in human adipocytes from functionally distinct adipose depots to elucidate epigenetic mechanisms underlying human obesity and obesity-induced metabolic disturbances. We discover extensive changes in DNA methylation associated with extreme obesity at epigenome-wide significance. Surprisingly, these methylation changes are largely adipose depot-specific with many more obesity-associated 5mC sites occurring in subcutaneous than visceral adipocytes. We surmise this depot specificity may be due to the local tissue microenvironment in the absence of technical, genetic or other known confounding factors.

By integrating our DNA methylation findings with adipocyte-specific transcriptomic and chromosomal interaction datasets, and cross-tissue enhancer-promoter catalogues, we statistically and functionally connect extreme obesity-associated 5mC sites to transcriptomic changes at >500 genes. These putative effector genes of obesity-associated methylation changes cluster in developmental, metabolic and inflammatory pathways, and encode proteins with key roles in adipogenesis, browning/beigeing of white adipocytes and insulin signalling. Of particular interest, we associate lower levels of DNA methylation in obese subcutaneous adipocytes with increased expression of a micro-RNA cluster, comprising MIR23A, MIR24-2 and MIR27A, whose members have been shown to inhibit *PPARG* signalling, suppress adipogenesis and induce insulin resistance^78–83^.

Our analyses co-localise extreme obesity-associated 5mC variations to functionally active genomic regions and transcription factor binding sites. They further suggest that TF activity may alter methylation status and that methylation status may impact TF activity at differentially methylated sites associated with obesity. These findings require experimental validation but are supported by existing evidence of reciprocal relationships between DNA methylation and TFs, and methylation-sensitivity of the TFs we identify^88–90, 93^. At a cellular level, many of the TF families linked to obesity-associated DNA methylation variations – AP1, KLF, SOX and ETS – have established or emergent roles in adipocyte biology and metabolism^64, 94–97, 102^, connecting disease-related DNA methylation variations to potential pathogenic pathways.

Contrary to most previous human obesity EWAS^22–24, 114–129^, we causally implicate up to 13% of extreme-obesity associated 5mC sites in obesity and obesity-induced metabolic disease susceptibility. Our MR findings should be regarded as causal estimates of the effects of methylation on phenotype under genetic control^130^. They do not indicate absence of causation at environmentally determined loci, the majority of our dataset. Complementary functional screens of target genes of causative methylation variations demonstrate novel effects on adipogenesis, *PPARG* signalling and adipocyte lipid handling, offering cellular mechanisms by which DNA methylation may promote obesity and its consequences.

New technologies for profiling DNA methylation and transcriptional regulation at single cell resolution will enable future studies to address the importance of cellular epigenetic heterogeneity, methylation and transcriptional dynamics, and spacial microenvironmental interactions, in obese adipose tissue remodelling^131, 132^. Nevertheless, we show that existing technologies that facilitate the study of epigenomic variations in larger numbers of individuals, and thus better capture human phenotypic diversity, remain a valuable tool for de-convoluting the epigenetic basis of human obesity and T2D. Combining these high-throughput approaches with precision epigenome tools^133–136^ will clarify the impact of DNA methylation, including at the sites we identify, on disease pathogenesis.

Taken together, our exploratory studies in human adipocytes begin to reveal genomic mechanisms and molecular signalling pathways through which DNA methylation may impact human obesity and its metabolic consequences. We provide new evidence of causation at a sizeable fraction of extreme obesity-associated 5mC sites, and a resource of epigenomic variations and genes for furthering our understanding of the human epigenome and its role in obesity and its metabolic complications.

## METHODS

### Study design

Case-control DNA methylation analyses were carried out in 95 subcutaneous and 95 visceral adipocyte samples from people with extreme-obesity and healthy controls in separate discovery and replication cohorts (**Supplementary Figure 1**). 44 participants in the discovery cohort and 42 participants in the replication cohort provided both subcutaneous and visceral adipocyte samples. The remaining subcutaneous and visceral adipocyte samples came from distinct individuals. RNA sequencing was carried out in adipocytes from the replication cohort.

### Participant selection and sample processing

Adipose tissue samples were obtained intraoperatively from morbidly obese individuals (mean (sd) BMI 44.8 (7.2) kg/m2) undergoing laparoscopic bariatric surgery and healthy controls (mean (sd) BMI 24.9 (3.3) kg/m2) undergoing non-bariatric laparoscopic abdominal surgery (**Supplementary Table 1**). Subcutaneous tissue was collected from abdominal surgical incision sites and visceral tissue from the omentum. Participants were unrelated, between 20-70 years of age, from a multi-ethnic background, and free from systemic illnesses not related to obesity. Controls and cases were well-matched for age (within 3-yrs), sex and ethnicity (**Supplementary Table 1**). People with treated T2D were excluded because of potential effects of hypoglycaemic medications on adipose tissue metabolism. All participants gave informed consent. The study was approved by the London — City Road and Hampstead Research Ethics Committee, United Kingdom (reference 13/LO/0477).

Whole tissue samples were processed immediately to isolate populations of primary adipocytes using established protocols^137, 138^. Polypropylene plastics were used to minimise adipocyte cell lysis. Tissues were cut into 1-2-mm3 pieces and washed in Hank’s buffered salt solution (HBSS), before digestion using type 1 collagenase (1mg/ml, Worthington) in a water bath at 37C shaking at 100-rpm for ∼30-min. Digested samples were filtered through a 300-micron nylon mesh to remove debris, and the filtered solution was centrifuged at low speed (500-g; 5-min; 4C). After removal of the oil layer, floating mature adipocytes were collected by pipette, washed in ∼5x volume of HBSS and recentrifuged. After 3 washes the clean adipocyte cell suspension was collected for snap freezing and storage at −80C.

### Quantification of DNA methylation

Genomic DNA and RNA were extracted in parallel from isolated adipocytes using the Qiagen AllPrep DNA/RNA/miRNA Universal Kit according to the manufacturer’s protocol for lipid-rich samples. Genome-wide DNA methylation was assayed using Illumina Infinium HumanMethylation450 (96 discovery samples) and EPIC (96 replication samples) beadchips. In both cohorts, all samples were randomised and processed in single batches. 0.2-0.5mcg of genomic DNA was bisulphite converted using EZ DNA Methylation-Direct Kits (Zymo Research, Irvine, CA). Bisulfite-treated DNA was denatured, neutralised and subjected to an overnight whole-genome amplification reaction. Amplified DNA was enzymatically fragmented, precipitated and resuspended for hybridisation to respective HumanMethylation450K and EPIC beadchips. After hybridization, beadchips were processed through a primer-extension protocol, stained, coated and then imaged using the HiScan System (Illumina).

Raw signal intensities were retrieved using the readIDAT function of the R package minfi (version 1.36.0, Bioconductor^139^), followed by background correction with the function bgcorrect.illumina from the same R package. Detection P values were derived using the function detectionP as the probability of the total signal (methylation + unmethylated) being detected above the background signal level, as estimated from negative control probes. Signals with detection P values ≥0.01 were removed. >99% of CG sites in both cohorts passed this quality control threshold. One visceral (discovery) and one subcutaneous (replication) sample with less than 95% of CG sites providing a detected signal were excluded. To reduce non-biological variability between observations, data were quantile normalized with the function normalizeQuantiles of the R package limma (version 2.12.0, Bioconductor). DNA methylation was quantified on a scale of 0-1, where 1 represents 100% methylation.

Separate principal component analyses (PCA) were carried out on HumanMethylation450K and EPIC positive control probe signal intensities. These probes assess multiple steps in the laboratory processing and the resulting principal components (PCs), which capture technical variability in the experiment, were included as covariates in our discovery and replication models to remove technical biases^140^.

### RNA sequencing

RNA sequencing was carried out in the 96 samples with paired DNA methylation results from the replication cohort. Sample order was randomised for library preparation and sequencing. Total RNA was quantified using the Qubit Fluorometer (RNA HS Assay) and aliquots of 10-ng were used for library preparation. RNA integrity numbers (RINs, Aligent 2100 Bioanalyzer) ranged from 2.5 to 7.9 (median 5.8), an indication of suboptimal transcript quality in isolated human adipocytes (stored at −80C). Sequencing libraries were therefore generated using the SMARTer Stranded Total RNA-Seq Kits (v2 Pico input) which uses Switching Mechanism at the 5’ end of RNA Template technology and random priming to make cDNA from low-quality, low-input RNA volumes. Takara Bio/Clontech recommendations were followed at each library preparation stage (fragmentation, first-strand cDNA synthesis, addition of adapters and indexing, AMPure bead purification, library amplification, AMPure bead repeat purification). Final libraries were validated using the Aligent BioAnalyzer (DNA1000 and High Sensitivity chips) and 2 failed libraries were removed. Sample-specific libraries were pooled at equimolar concentrations (4nM) to avoid batch effects and sequenced according to the manufacturer’s protocols using the HiSeq 4000 (16 lanes). Each sample was sequenced to >80 million 100-bp paired-end reads.

Raw sequencing data was demultiplexed using Bcl2Fastq (version 2.20) and read quality was checked using fastQC (version 0.11.5). Reads were adapter trimmed (Illumina generic adapters) and mapped against the Ensembl human reference genome GRCh37 v87 using the splice aware aligner STAR (version 2.6.0, options: -outFilterMultimapNmax 20 -outFilterMismatchNoverReadLmax 0.04). Lane level bams were merged, sorted and indexed using Samtools (version 1.9) to create library level bams for each sample. Mapping rates and quality control measures were evaluated level using Picard (version 2.18.12) and RseQC (version 2.6.4), and summarised using MultiQC (version 1.9). Raw counts per gene were generated using FeatureCounts (subread version 1.6.2); multimapped reads were included (options: -M -fraction) to capture short reads resulting from lower quality transcripts. 2 samples with low number of aligned reads (<15M) reflecting low-quality libraries were excluded. 3 other outlying samples with low median transcript integrity (TIN) or evidence of GC bias were removed (**Supplementary Figure 10A and 10B**). After exclusions, gene expression results were available for 43 subcutaneous and 46 visceral adipocyte samples and 24,187 genes (counts ≥5 in ≥20% of samples). As a final QC step, surrogate variable analysis (SVA^141^) was carried out on variance stabilising transformed counts (generated using DESeq2, version 1.28.0^142^) for each sample to capture unwanted variation in our RNA sequencing data, particularly that arising due to differences in transcript and sequencing quality (**Supplementary Figure 10C**). The resulting surrogate variables (SV) were included as covariates in 5mC regression models to remove associations driven by sample and sequencing biases.

### Epigenome-wide association analyses

All methylation-phenotype analyses were carried out separately in subcutaneous and visceral adipocytes. 44,101 probes were removed because of known cross hybridisation^143, 144^ or presence of a common genetic variant (SNP, indels, or structural variation, 1000G European phase 3 dataset, MAF>1%) within the probe sequence. After QC and removal of problem probes, only the 401,595 single CG sites present on both the Illumina HumanMethylation450 and EPIC beadchips were investigated. Single markers passing quality control were tested for association with extreme obesity using linear regression and an established analytic strategy to reduce batch and other technical confounding effects^145^. Obesity status was used as the predictor with methylation as the outcome variable to generate %-methylation differences between cases and controls. Covariate adjustments were made for potential biological (age, sex and ethnicity) and technical confounding variables (the first 4 control probe PCs which explained >95% of control probe technical variation). Association results from the discovery and replication cohorts were combined using inverse variance weighted (IVW) meta-analysis and evaluated for heterogeneity.

Statistical significance was inferred at FDR<0.01^33, 34^ in the discovery sample to provide an inclusive set of biologically relevant results. A more stringent cut off of (i) FDR<0.01 in the replication sample (with consistent direction of effect) and (ii) combined P<1×10-7 (epigenome-wide significance, EWS^140^) in the combined discovery and replication samples was then used to define significant associations. Markers associated with obesity at P<1×10-7 within ±5000-bp of each other were considered as a single genomic locus. A ±5000-bp distance was selected to take into account the known sizes of discrete regions of DNA methylation, for example CGIs^146^. At each independent locus the CG site with lowest P value for association with obesity was defined as the sentinel marker. An exact binomial test (R function *binom.test*) was used to test whether consist directions of effect between discovery and replication, and between subcutaneous and visceral, were observed more often than expected by chance.

### Genetic confounding and adipocyte purity

Genotype data for each participant was generated from whole blood using Illumina Infinium OmniExpress-24 v1.2 beadchips. We removed directly genotyped SNPs with call rates <90%, minor allele frequency <0.01, Hardy-Weinberg Equilibrium P<1×10-6, SNPs on sex chromosomes and duplicated SNPs. After quality control, 649,007 SNPs were taken forward for imputation. SHAPEIT^147^ was used to infer haplotypes, and imputation was carried out in IMPUTE2 (version 2.3.2^148^) using the 1000 genome reference panel Phase 3 (all ancestries). Each chromosome was divided into 5Mb chunks for imputation and merged at the end. A random seed was supplied automatically. Effective population size (Ne) reflecting genetic diversity was 20,000 as recommended when using a multi-population reference panel. After imputation, genotype data was available for 81,656,368 SNPs. Sensitivity analyses were carried out in combined discovery and replication results (IVW meta-analysis). Multivariate regression models with and without genetic confounding factors were compared (**Supplementary Figures 2A and 2B**). First, PCA was performed on participant GWAS data and the first 5 PCs explaining >95% of inter-individual variation were included as covariates to adjust for population stratification. Second, the effects of cis-SNPs (within 500-kb) on methylation-phenotype associations were examined by including (i) the cis-SNP most strongly associated with DNA methylation and (ii) all independent cis-SNPs associated with DNA methylation at FDR<0.01 (pairwise LD R2<0.8) in multivariate regression models. Gaphunting from the Minfi package was also used to identify and flag methylation sites at which the distribution of methylation was consistent with an underlying SNP driven effect (version 1.36.0^36, 139^ **Supplementary Tables 2A and 2B**).

Purity of the isolated adipocytes was evaluated using two approaches. First, in the replication cohort, RNA sequencing results for immune and stromovascular cell-specific genes were used to evaluate associations driven by potential contamination (broad immune cell – CD45/PTPRC; monocyte/macrophage – CD68, CD11b/ITGAM, CD14; B/T cell – TNFRSF8, CD19, CD4, CD8A, CD2; NK cell – NCAM1; endothelial cell – CD31; preadipocyte – DLK1/PREF1, CD34). Replication models without and with variance stabilised transformed gene expression counts (DESeq2, version 1.28.0) were compared for each immune-or stromovascular cell-specific gene (**Supplementary Figures 3A and 3B**). PCA analysis was also carried out to summarise the variance across the potential contaminating cell genes, and models without and with PCs were compared (**Supplementary Figures 3A and 3B**). Second, in the combined discovery and replication cohorts, SVA analysis was performed to capture unexplained variation due to cellular heterogeneity (resulting from contamination) and other potential confounding factors in our high-dimensional DNA methylation data^141, 149–151^. The resulting SV were then included as covariates in regression models to test whether the observed associations were driven by impurity or other unknown confounding factors (**Supplementary Figure 4**).

### Methylation-expression analyses

Individual sentinel 5mC sites were assigned to potential target genes using three approaches. First, sentinels in exons, 5/3’ UTRs or within 5-kb of a promoter were assigned to overlapping genes. A 5-kb cut-off to define a promoter region was selected based on an observed drop off in the number of 5mC sentinels beyond this distance from their respective TSS (using sequential 1-kb bins). Second, for 5mC sites in intronic/intergenic regions, we overlapped the sentinel methylation site with distal chromosomal intervals connected to proximal gene transcription start sites from: i. Adipocyte Capture Hi-C results; and ii. the GeneHancer enhancer-promoter inference database^42^. Human adipocyte Capture Hi–C data was available at GEO (Accession ID: GSE110619^55^) as pre-processed and pre-called interactions. GeneHancer interactions were taken from the combined standard and elite functional interaction sets, with removal of standard interactions defined by proximity alone. Third, for those intronic/intergenic sentinels not assigned to a functional targeted gene, we intersected the sentinel sites with adipocyte-specific TADs, and took all genes within intersected sentinel-TAD pairs as potential targets. Adipocyte TADs were called from available Hi-C data generated at day 3 of human adipocyte differentiation in vitro (GEO, Accession ID: GSE109924^56^). TADs were called from bed files, using Arrowhead at 25-kb resolution with default parameters, and merged, removing hierarchal structures by retaining largest domains to maximise TAD coverage, for a final set of 5323 domains of median 425-kb, range 125-kb to 4,025-kb intervals.

Sentinel 5mC sites were tested for association with expression of each of their functionally assigned target genes using linear regression. Initially, we carried out association testing separately in subcutaneous and visceral adipocytes, but had limited power to detect methylation-expression associations. Thus, for our final analyses we used the combined subcutaneous and visceral adipocyte datasets to identify methylation-expression relationships, and mixed-effect modelling to control for sample relatedness (i.e. subcutaneous and visceral adipocytes from the same individuals). Mixed effect models were implemented using Dream analysis from the Bioconductor package variancePartition (version 1.18.0) in R; methylation betas were used as the predictor and logCPM transformed gene expression counts (voomWithDreamWeights) as the outcome. Study participant ID was included as a random effect. Two methylation control probe PCs, two SVs generated using SVA of the gene expression counts, age, sex, and ethnicity, and RNA integrity numbers (RINs) were included as fixed effects to adjust for technical, biological and sample-related biases. Mixed model results from Dream were compared with those from the nlme R package (version 3.1-149, using variance stabilised transformed counts from DESeq2, version 1.28.0) as a further sensitivity test. Statistical significance was inferred at FDR<0.01 (qvalue version 2.20.0^33^) based on the number of methylation-target gene pairs analysed.

### Functional enrichment analyses

All functional enrichment analyses were carried out using permutation testing as Illumina methylation arrays preferentially evaluate pre-selected genomic sites (e.g. CG islands) and well-annotated genes. For each sentinel CG site, we identified a permutation set of 1000 unique CG sites with equivalent methylation levels and variability in their respective subcutaneous or visceral adipocyte samples, using the following criteria: i. difference in mean between sentinel and permutation; ii. difference in standard deviation (sd) between sentinel and permutation; and iii. >5-kb distance between sentinel and permutation (i.e. not at the same genomic locus). For each sentinel, difference in mean and sd were based on a sliding scale starting at mean 0.025 and sd 0.0025 and increasing incrementally by mean 0.025 and sd 0.0025 until >1000 independent permutated CG sites were achieved. This approach was selected because a low fixed mean/sd was too stringent to generate 1000 permutations at some sentinels, while a higher fixed mean/sd was too permissive at other sentinels.

For genomic enrichment analyses, we compared the number of sentinels located in a genomic feature (observed frequency) with that in each of the 1000 permutation sets (expected frequency). For human obesity and metabolic disease GWAS enrichment analyses, we identified methylation sites and GWAS SNPs (P<5×10-8) in shared same Adipocyte Roadmap Chromatin State (E025), and compared the frequencies between the sentinels and the permuted background. We used the same permutation-based approach for our nearest gene pathway analyses, but limited our analyses to one sentinel per gene and one permutation per gene, to avoid recounting genes. The nearest gene to each sentinel was identified using the ChIPseeker annotation package (version 1.28.3^152^), which prioritises overlaps in promoters over other features (ranking: promoter, 5/3’UTR, Exon, Intron, Intergenic). Nearest genes were cross-referenced with the Molecular Signatures Database (MsigDB, hallmark, curated and ontology gene sets^153, 154^). Differences in observed and expected frequencies were calculated using the Fishers Exact test and Empirical P values. Gene set enrichment analyses of genes associated with methylation in adipocytes were carried out in gProfiler^87^, using a background of the nearest gene to each of the 1000 permuted CG sites, limited to genes expressed in our human adipocyte RNA sequencing data.

### Transcription factor binding site analyses

De novo transcription factor binding motif enrichment analysis was performed using the script fingMotifGenome.pl from the Homer program package (version 4.11.1). Subcutaneous and visceral sentinels were investigated separately. Regions of interest were selected by extending each sentinel site for ±150-bp on each side. The enrichment analysis was done using two different backgrounds as controls: i. the ±150-bp regions around the 1000 permutation sites specific to each sentinel; and ii. genomic regions with GC% matching those of sentinel regions. At each motif, we identified the 10 transcription factors most likely to interact with that motif (best match between known motifs and the discovered motif) to provide a sizeable but manageable number of TFs. We then restricted these TFs to those expressed in our human RNA sequencing data (counts ≥5 in ≥20% of samples).

Positions in the region around the sentinel of respective motifs for analysis were inferred with the annotatePeaks.pl script from the Homer program package, with the options -rmrevopp to account for palindromic motifs. Genomic CpG sites relative to motifs were retrieved using the Bioconductor package seqPattern (version 1.20.0) in R. Correlation analyses of TF expression and methylation of their corresponding sentinels were carried out in the depot in which the sentinel was identified. Relationships between TFs, their respective sentinels and the predicted target genes of each sentinel (assigned sequentially first by promoter/exon/UTR overlap, then by functional interaction, then by shared TAD) were studied in combined subcutaneous and visceral adipocyte samples to increase power. Associations between TF and target gene expression were examined (i) without and (ii) with adjustment for sentinel DNA methylation level to explore the effects of sentinel methylation sites on TF-target gene relationships. Mixed effects models were carried out in the package nlme package (version 3.1_143) to adjust for sample relatedness; Age, Sex, Ethnicity, two methylation control probe PCs and two gene expression SVs were included as fixed effect covariates to adjust for potential confounding variables. For all TF expression analyses, variance stabilising transformed counts (DESeq2, version 1.28.0) were used.

### Genetic association analyses

Two sample Mendelian Randomisation (MR) analyses were carried out to investigate causal relationships between individual sentinel 5mC sites and human obesity phenotypes using: i. 588 whole subcutaneous adipose tissue (WSAT) samples from the Twins UK cohort; and ii. summary results from recent large-scale human GWAS.

The Twins UK cohort is a nationwide registry of healthy volunteer twins in the United Kingdom, with about 14,000 registered twins since 1992, predominately Caucasian female (84%) and equal number of monozygotic and same-sex dizygotic twins. Twins UK phenotypic measurements, adipose tissue biopsies, genome-wide SNP and DNA methylation assays were performed as previously described^104–106^. Briefly, samples were genotyped using HumanHap300, HumanHap610Q, HumanHap1M Duo, and HumanHap1.2M Duo 1M arrays. Haplotypes from IMPUTE2 (without a reference panel) were used for fast imputation to the 1000 Genomes phase 1 dataset. Imputed SNPs were excluded based on Hardy Weinberg equilibrium (P<1e-6), allele frequency cut-offs (MAF<0.01), missingness (>5%) and imputation quality (info score<0.8). Individuals with mis-assigned sex or ancestry outliers were removed. Ancestry outliers (>7 SD) were obtained from PLINK 2.0 (unrelated) and GENESIS (related participants). Related individuals with IBS > 0.125 (PLINK 2.0) were also excluded. DNA methylation profiles in adipose tissue biopsies were obtained as described previously^106^. Methylation results were available for 588 out of 596 twins after further quality control analyses^107^. All individuals were female (mean (sd) age 59.1 (9.4)).

Human GWAS comprised: BMI as a measure of obesity (GIANT 2018, transethnic^47^); WHR adjusted for BMI as a measure of central adiposity (GIANT 2018, transethnic^48^); fasting glucose and insulin (MAGIC, Europeans, unpublished); HbA1c (MAGIC 2017^108^); T2D and T2D adjusted from BMI (DIAGRAM 2018, Europeans^49^).

Cis-SNPs within ±500-kb of each subcutaneous and visceral sentinel were tested for association with change in sentinel DNA methylation level in WSAT using linear regression. DNA methylation levels were adjusted for technical covariates, age, predicted smoking, family relatedness, genetic principal components (PCs) and non-genetic DNA methylation PCs. Methylation-genotype associations were evaluated in the MatrixEQTL package in R (version 2.1.0) using linear models, with the adjusted methylation values as the dependent variable and the dosage of alternative allele the independent variable. Ambiguous palindromic cis-SNPs with MAF>0.42 were removed. For each sentinel, cis-SNPs were clumped (linkage disequilibrium (LD) R2<0.01) and independent methylation quantitative trait locus (mQTL) SNPs associated with DNA methylation at P<0.05 (Bonferroni corrected for the number of SNPs) were selected. Primary MR analyses of these mQTL SNPs and human GWAS phenotypes were implemented in R using the TwoSampleMR package (version 0.5.1^109, 110^). Causal relationships were tested using the most powerful MR method (Wald Test for single mQTL SNPs, and Inverse Variance Weighted method for multiple mQTL SNPs). Cause-consequence directions of effect were evaluated using the Steiger directionality test, which compares SNP-methylation and SNP-phenotype R2 values. Potential causal effects of methylation on phenotype inferred if both the MR and Steiger tests passed a significance threshold of FDR<0.01.

MR sensitivity testing was carried out using 2 approaches. First, we evaluated MR assumptions by repeating our MR analyses using correlated cis-SNPs (within ±500-kb, clumped at LD R2 >0.8, and associated with methylation at P<0.05 Bonferroni corrected) in the R package MendelianRandomization (version 0.4.1); correlated cis-SNPs were used as no sentinels had ≥3 uncorrelated cis-SNPs for such analyses. MR IVW and MR Egger regression were used to test for: i. replication; ii. heterogeneity between SNPs; and iii. evidence of horizontal pleiotropy; at each sentinel with ≥3 correlated cis-mQTLs. Second, we replication tested WSAT mQTLs implicated in positive MR results amongst our subcutaneous and visceral adipocyte samples using SNP as the predictor, methylation beta as the outcome, and adjusting for biological and technical confounders (age, sex, ethnicity, control probe PCs 1-4). For WSAT mQTL SNPs not present in our adipocyte dataset, we identified a proxy SNP present in adipocytes (the cis-SNP (±500-kb) with the greatest pairwise LD with the mQTL SNP and minimum R2>0.8) and used the correlated allele to evaluate for association with methylation and concordance of directions of effect.

### In vitro studies

The 3T3-L1 pre-adipocyte mouse cell line (ATCC-CL-173) was obtained commercially (LGC). Pre-adipocytes were grown in Dulbecco’s Modified Eagles Media with 10% newborn calf serum (NCS) and 1% penicillin/ streptomycin (P/S). Two days post-confluence (Day 0) differentiation was induced by switching cells to DMEM supplemented with 10% foetal bovine serum (FBS), 10-µg/ml insulin, 0.5-mM IBMX, 1-µM dexamethasone and 2-µM rosiglitazone. On day 2, cell media was refreshed with insulin media (DMEM containing 10-µg/ml insulin). Cells were maintained at 37C and 5% CO2 and were differentiated in 10-cm dishes until undergoing siRNA reverse transfection.

Early differentiation 3T3-L1 adipocytes were reverse transfected at Day 2 of differentiation with Silencer Select siRNA’s (Ambion) as described below, and similar to that described previously^155^. Two different siRNAs against each target were used in concert to enhance target gene knockdown. Cells were transfected with siRNA’s against either *Limd2* (ID; s85672; s85674), *Prrc2a* (ID; s79278; s79279) or a non-silencing (NS) (ID; Negative control #1) siRNA. RNAiMax lipofectamine transfection reagent (Life Technologies) and siRNAs were diluted separately in Opti-MEM media (Gibco), mixed together, added to empty wells and incubated for 20-mins before cell suspension was seeded. To each well of a 6-well and 12-well plate, 50-pmol of siRNA (25-pmol each siRNA) and 30-pmol siRNA (15-pmol each siRNA) were added respectively. For NS control, a single siRNA was used at the same total pmol quantity as for targets. Differentiating 3T3-L1 adipocytes in 10-cm dishes were detached at Day 2 of differentiation by treating with Trypl Express (Gibco) for 10-mins. Cells were counted, resuspended at 450,000 cells/ml in DMEM insulin media (insulin, 10-µg/ml), and added to 12-well (1ml; 450,000 cells/well) and 6-well plates (2ml; 900,000 cells/well) containing the pre-incubated siRNA transfection mix. The next day cells were refreshed with new insulin media (10-µg/ml insulin). On day 6, siRNA treated differentiated cells were harvested for RNA (6-well plates) or assayed for lipid accumulation (12-well plate; Oil Red O).

Oil red O (ORO) staining was performed to assess lipid accumulation in mature 3T3-L1 cells (Day 6) that were reverse transfected with siRNA at early differentiation (Day 2). The protocol used was similar to that described previously^156^ with modifications. Cells were washed with PBS and fixed with 10% neutral formalin for 1-hr. After formalin removal, cells were washed with sterile water then exposed to 60% isopropanol for 3-mins. After removal of 60% isopropanol, cells were stained with ORO solution (Sigma) for 10-mins, and then washed with water until all extracellular ORO was completely removed. At this point images of ORO staining at 4X and 10X magnification were acquired using the Evos m7000 microscope (Thermo Scientific). Cells were treated with 100% isopropanol for 10-mins to extract ORO stain from lipid in cells for quantification by measuring elute absorbance at 500-nM using SpectrumMax 340PC plate reader (Molecular Devices). Samples were added to 96-well plate in quadruplicate along with known ORO quantities (µg/ml) to make a standard curve to calculate µg of ORO eluted. Following ORO elution, cells were washed 2x with water to allow crystal violet (CV) nuclear staining for relative cell number normalisation. Cells were stained with 0.05% CV for 10-mins, followed by 4x 10-min washes with water to remove all extracellular CV. SDS (1%) was added to cell plates and incubated for 10-mins with constant orbital agitation at 150rpm to lyse cells and allow CV absorbance in the lysate to be measured at 560-nM using the SpectrumMax 340PC plate reader. CV samples were added to 96-well plates in quadruplicate along with known CV quantities (µg/ml) to generate a standard curve to calculate µg of CV and to thus normalise ORO data to relative cell number.

Total RNA was isolated from siRNA transfected 3T3-L1 adipocytes at Day 6 using Qiazol reagent and the RNeasy mini kit (Qiagen) according to manufacturers’ instructions, with on-column DNase (Qiagen) treatment performed during RNA isolation. The High-Capacity RNA-to-cDNA kit was used to generate cDNA by reverse transcription of 1-µg total RNA. RT-qPCR gene expression analysis was performed using the CFX384 Touch Real-Time PCR Detection System (BioRad), SSO advanced Universal SYBR Green Supermix mix, gene-specific primers (500-nM final concentration) and cDNA in a 10-µl total reaction volume. qPCR conditions were: 3-mins at 95C, then 40 cycles of 95C for 10-secs, 60C for 30-secs and followed by melting curve analysis from 65-95C in 0.5C steps at 5 secs/step. Sequences of primers used in qPCR analysis are listed in **Supplementary Table 19**. Gene expression was quantified using the delta-delta Ct (2-ΔΔCT) method and is shown relative to the non-silencing group. Two housekeeping genes *Nono* and *Ywhaz* were utilised, with their geometric mean expression being used for normalisation. Effects of knockdown on genes involved in adipocyte differentiation (*Pparg*), insulin signalling (*Glut4*, *Irs1*), lipid uptake (*Lpl*), lipid storage (*Fasn*, *Acaca*, *Scd1*) and lipid mobilisation (*Fabp4*, *Hsl*) were evaluated^157^.

GraphPad Prism was used to perform Student’s t-tests used in analyses of gene expression and Oil Red O. All data shown relative to non-silencing group and presented as means ± SEM.

## Data Availability

All data produced in the present study are available upon reasonable request to the authors, and will be made openly available after critical review.

## ACKNOWLEDGEMENTS

This work was funded by the Medical Research Council UK (MR/K002414/1), the Wellcome Trust (219602/Z/19/Z) and the National Institute for Health Research (NIHR) Imperial Biomedical Research Centre (BRC). The Illumina HumanMethylation450 Beadchip data used in this research was generated by the High-Throughput Genomics Group at the Wellcome Trust Centre for Human Genetics (funded by Wellcome Trust grant reference 090532/Z/09/Z). The Illumina EPIC Beadchip data were generated in collaboration with RS. The NIHR Imperial BRC Genomics Facility provided resources and support that contributed to the reported RNA sequencing results. The TwinsUK research components were funded by JPI ERA-HDHL DIMENSION via the Biotechnology and Biological Sciences Research Council (BB/S020845/1). TwinsUK is funded by the Wellcome Trust, Medical Research Council, European Union, Chronic Disease Research Foundation (CDRF), Zoe Global Ltd and the NIHR-funded BioResource, Clinical Research Facility and Biomedical Research Centre based at Guy’s and St Thomas’ NHS Foundation Trust in partnership with King’s College London. We thank the participants and research staff who made this possible.

## REFERENCES

1. WHO. World Health Organisation Fact Sheets. Obesity and Overweight. https://www.who.int/news-room/fact-sheets/detail/obesity-and-overweight (2021).

2. Collaborators, G. 2015 O. et al. Health Effects of Overweight and Obesity in 195 Countries over 25 Years. New Engl J Medicine 377, 13–27 (2017).

3. Blüher, M. Obesity: global epidemiology and pathogenesis. Nat Rev Endocrinol 15, 288–298 (2019).

4. González-Muniesa, P., et al. Obesity. Nat Rev Dis Primers 3, 17034 (2017).

5. Reilly, S. M. & Saltiel, A. R. Adapting to obesity with adipose tissue inflammation. Nat Rev Endocrinol 13, 633–643 (2017).

6. Srivastava, G. & Apovian, C. M. Current pharmacotherapy for obesity. Nat Rev Endocrinol 14, 12– 24 (2018).

7. Ling, C. & Rönn, T. Epigenetics in Human Obesity and Type 2 Diabetes. Cell Metab 29, 1028–1044 (2019).

8. Loh, M., Zhou, L., Ng, H. K. & Chambers, J. C. Epigenetic disturbances in obesity and diabetes: Epidemiological and functional insights. Mol Metab 27, S33–S41 (2019).

9. Greenberg, M. V. C. & Bourc’his, D. The diverse roles of DNA methylation in mammalian development and disease. Nat Rev Mol Cell Bio 20, 590–607 (2019).

10. Fernandez-Twinn, D. S., Hjort, L., Novakovic, B., Ozanne, S. E. & Saffery, R. Intrauterine programming of obesity and type 2 diabetes. Diabetologia 62, 1789–1801 (2019).

11. Chen, Z. et al. DNA methylation mediates development of HbA1c-associated complications in type 1 diabetes. Nat Metabolism 2, 744–762 (2020).

12. Chen, R. et al. Longitudinal personal DNA methylome dynamics in a human with a chronic condition. Nat Med 24, 1930–1939 (2018).

13. Xue, A. et al. Genome-wide association analyses identify 143 risk variants and putative regulatory mechanisms for type 2 diabetes. Nat Commun 9, 2941 (2018).

14. Quach, A. et al. Epigenetic clock analysis of diet, exercise, education, and lifestyle factors. Aging 9, 419–446 (2017).

15. Bacos, K. et al. Blood-based biomarkers of age-associated epigenetic changes in human islets associate with insulin secretion and diabetes. Nat Commun 7, 11089 (2016).

16. Boškovi, A. & Rando, O. J. Transgenerational Epigenetic Inheritance. Annu Rev Genet 52, 21–41 (2018). ^ć^

17. Rakyan, V. K., Down, T. A., Balding, D. J. & Beck, S. Epigenome-wide association studies for common human diseases. Nat Rev Genet 12, 529–541 (2011).

18. Birney, E., Smith, G. D. & Greally, J. M. Epigenome-wide Association Studies and the Interpretation of Disease -Omics. Plos Genet 12, e1006105 (2016).

19. Lappalainen, T. & Greally, J. M. Associating cellular epigenetic models with human phenotypes. Nat Rev Genet 18, 441–451 (2017).

20. Stricker, S. H., Köferle, A. & Beck, S. From profiles to function in epigenomics. Nat Rev Genet 18, 51–66 (2017).

21. Teschendorff, A. E. & Relton, C. L. Statistical and integrative system-level analysis of DNA methylation data. Nat Rev Genet 19, 129–147 (2018).

22. Banos, D. T. et al. Bayesian reassessment of the epigenetic architecture of complex traits. Nat Commun 11, 2865 (2020).

23. Wahl, S. et al. Epigenome-wide association study of body mass index, and the adverse outcomes of adiposity. Nature 541, 81–86 (2017).

24. Dahlman, I. et al. The fat cell epigenetic signature in post-obese women is characterized by global hypomethylation and differential DNA methylation of adipogenesis genes. Int J Obesity 39, 910–919 (2015).

25. Bradford, S. T. et al. Methylome and transcriptome maps of human visceral and subcutaneous adipocytes reveal key epigenetic differences at developmental genes. Sci Rep-uk 9, 9511 (2019).

26. Paul, D. S. et al. Increased DNA methylation variability in type 1 diabetes across three immune effector cell types. Nat Commun 7, 13555 (2016).

27. Rosen, E. D. & Spiegelman, B. M. What We Talk About When We Talk About Fat. Cell 156, 20– 44 (2014).

28. Kahn, C. R., Wang, G. & Lee, K. Y. Altered adipose tissue and adipocyte function in the pathogenesis of metabolic syndrome. J Clin Invest 129, 3990–4000 (2019).

29. Tchkonia, T. et al. Mechanisms and Metabolic Implications of Regional Differences among Fat Depots. Cell Metab 17, 644–656 (2013).

30. Schleinitz, D. et al. Identification of distinct transcriptome signatures of human adipose tissue from fifteen depots. Eur J Hum Genet 28, 1714–1725 (2020).

31. You, D. et al. Dnmt3a is an epigenetic mediator of adipose insulin resistance. Elife 6, e30766 (2017).

32. Villivalam, S. D. et al. TET1 is a beige adipocyte-selective epigenetic suppressor of thermogenesis. Nat Commun 11, 4313 (2020).

33. Storey, J. D. & Tibshirani, R. Statistical significance for genomewide studies. Proc National Acad Sci 100, 9440–9445 (2003).

34. Wasserstein, R. L. & Lazar, N. A. The ASA’s Statement on p-Values: Context, Process, and Purpose. Am Statistician 70, 129–133 (2016).

35. Crujeiras, A. B. et al. DNA methylation map in circulating leukocytes mirrors subcutaneous adipose tissue methylation pattern: a genome-wide analysis from non-obese and obese patients. Sci Rep-uk 7, 41903 (2017).

36. Andrews, S. V., Ladd-Acosta, C., Feinberg, A. P., Hansen, K. D. & Fallin, M. D. “Gap hunting” to characterize clustered probe signals in Illumina methylation array data. Epigenet Chromatin 9, 56 (2016).

37. Jones, P. A. Functions of DNA methylation: islands, start sites, gene bodies and beyond. Nat Rev Genet 13, 484–492 (2012).

38. Schübeler, D. Function and information content of DNA methylation. Nature 517, 321–326 (2015).

39. Consortium, T. F. et al. An atlas of active enhancers across human cell types and tissues. Nature 507, 455–461 (2014).

40. Ienasescu, H. et al. On-the-fly selection of cell-specific enhancers, genes, miRNAs and proteins across the human body using SlideBase. Database 2016, baw144 (2016).

41. Kundaje, A. et al. Integrative analysis of 111 reference human epigenomes. Nature 518, 317–330 (2015).

42. Fishilevich, S. et al. GeneHancer: genome-wide integration of enhancers and target genes in GeneCards. Database 2017, bax028 (2017).

43. Ziller, M. J. et al. Charting a dynamic DNA methylation landscape of the human genome. Nature 500, 477–481 (2013).

44. Schultz, M. D. et al. Human body epigenome maps reveal noncanonical DNA methylation variation. Nature 523, 212–216 (2015).

45. Li, P. et al. Epigenetic dysregulation of enhancers in neurons is associated with Alzheimer’s disease pathology and cognitive symptoms. Nat Commun 10, 2246 (2019).

46. Song, Y. et al. Dynamic Enhancer DNA Methylation as Basis for Transcriptional and Cellular Heterogeneity of ESCs. Mol Cell 75, 905–920.e6 (2019).

47. Yengo, L. et al. Meta-analysis of genome-wide association studies for height and body mass index in ∼700000 individuals of European ancestry. Hum Mol Genet 27, 3641–3649 (2018).

48. Pulit, S. L. et al. Meta-analysis of genome-wide association studies for body fat distribution in 694 649 individuals of European ancestry. Hum Mol Genet 28, 166–174 (2019).

49. Mahajan, A. et al. Fine-mapping type 2 diabetes loci to single-variant resolution using high-density imputation and islet-specific epigenome maps. Nat Genet 50, 1505–1513 (2018).

50. Study, T. L. C. et al. Genetic studies of body mass index yield new insights for obesity biology. Nature 518, 197–206 (2015).

51. Heard-Costa, N. L. et al. NRXN3 Is a Novel Locus for Waist Circumference: A Genome-Wide Association Study from the CHARGE Consortium. Plos Genet 5, e1000539 (2009).

52. Lee, K. Y. et al. Tbx15 Defines a Glycolytic Subpopulation and White Adipocyte Heterogeneity. Diabetes 66, 2822–2829 (2017).

53. Consortium, T. Adipog., et al. New genetic loci link adipose and insulin biology to body fat distribution. Nature 518, 187–196 (2015).

54. Grant, S. F. A. The TCF7L2 Locus: A Genetic Window Into the Pathogenesis of Type 1 and Type 2 Diabetes. Diabetes Care 42, 1624–1629 (2019).

55. Pan, D. Z. et al. Integration of human adipocyte chromosomal interactions with adipose gene expression prioritizes obesity-related genes from GWAS. Nat Commun 9, 1512 (2018).

56. Paulsen, J. et al. Long-range interactions between topologically associating domains shape the four-dimensional genome during differentiation. Nat Genet 51, 835–843 (2019).

57. Boucher, J., Kleinridders, A. & Kahn, C. R. Insulin receptor signaling in normal and insulin-resistant states. Csh Perspect Biol 6, a009191–a009191 (2014).

58. Kadowaki, T. et al. Adiponectin and adiponectin receptors in insulin resistance, diabetes, and the metabolic syndrome. J Clin Invest 116, 1784–1792 (2006).

59. Gallardo-Montejano, V. I. et al. Perilipin 5 links mitochondrial uncoupled respiration in brown fat to healthy white fat remodeling and systemic glucose tolerance. Nat Commun 12, 3320 (2021).

60. Lee, S.-M. et al. FABP3-mediated membrane lipid saturation alters fluidity and induces ER stress in skeletal muscle with aging. Nat Commun 11, 5661 (2020).

61. Shi, H., Cave, B., Inouye, K., Bjorbaek, C. & Flier, J. S. Overexpression of Suppressor of Cytokine Signaling 3 in Adipose Tissue Causes Local but Not Systemic Insulin Resistance. Diabetes 55, 699– 707 (2006).

62. Kilicarslan, M. et al. RBP4 increases lipolysis in human adipocytes and is associated with increased lipolysis and hepatic insulin resistance in obese women. Faseb J 34, 6099–6110 (2020).

63. Ding, L., et al. Akt3 inhibits adipogenesis and protects from diet-induced obesity via WNK1/SGK1 signaling. Jci Insight 2, e95687 (2017).

64. Wu, Z. & Wang, S. Role of kruppel-like transcription factors in adipogenesis. Dev Biol 373, 235– 243 (2013).

65. Hilgendorf, K. I. et al. Omega-3 Fatty Acids Activate Ciliary FFAR4 to Control Adipogenesis. Cell 179, 1289–1305.e21 (2019).

66. Ramalingam, L., Oh, E. & Thurmond, D. C. Doc2b enrichment enhances glucose homeostasis in mice via potentiation of insulin secretion and peripheral insulin sensitivity. Diabetologia 57, 1476– 1484 (2014).

67. Li, Y. et al. Thioesterase superfamily member 1 undergoes stimulus-coupled conformational reorganization to regulate metabolism in mice. Nat Commun 12, 3493 (2021).

68. Xiao, Y. et al. Deficiency of PRKD2 triggers hyperinsulinemia and metabolic disorders. Nat Commun 9, 2015 (2018).

69. Song, W., et al. Organic cation transporter 3 (Oct3) is a distinct catecholamines clearance route in adipocytes mediating the beiging of white adipose tissue. Plos Biol 17, e2006571 (2019).

70. Chen, Q. et al. SIRT6 Is Essential for Adipocyte Differentiation by Regulating Mitotic Clonal Expansion. Cell Reports 18, 3155–3166 (2017).

71. Naeve, G. S. et al. Neuritin: A gene induced by neural activity and neurotrophins promotes neuritogenesis. Proc National Acad Sci 94, 2648–2653 (1997).

72. Pasterkamp, R. J. Getting neural circuits into shape with semaphorins. Nat Rev Neurosci 13, 605– 618 (2012).

73. Südhof, T. C. Synaptic Neurexin Complexes: A Molecular Code for the Logic of Neural Circuits. Cell 171, 745–769 (2017).

74. Cohen, P. et al. Ablation of PRDM16 and Beige Adipose Causes Metabolic Dysfunction and a Subcutaneous to Visceral Fat Switch. Cell 156, 304–316 (2014).

75. Stine, R. R. et al. EBF2 promotes the recruitment of beige adipocytes in white adipose tissue. Mol Metab 5, 57–65 (2016).

76. Sun, Y. et al. FGF9 inhibits browning program of white adipocytes and associates with human obesity. J Mol Endocrinol 62, 79–90 (2019).

77. Rajbhandari, P. et al. IL-10 Signaling Remodels Adipose Chromatin Architecture to Limit Thermogenesis and Energy Expenditure. Cell 172, 218–233.e17 (2018).

78. Yao, F. et al. Adipogenic miR-27a in adipose tissue upregulates macrophage activation via inhibiting PPARγ of insulin resistance induced by high-fat diet-associated obesity. Exp Cell Res 355, 105–112 (2017).

79. Lozano-Bartolomé, J. et al. Altered Expression of miR-181a-5p and miR-23a-3p Is Associated With Obesity and TNFα-Induced Insulin Resistance. J Clin Endocrinol Metabolism 103, 1447–1458 (2018).

80. Jin, M. et al. MicroRNA-24 promotes 3T3-L1 adipocyte differentiation by directly targeting the MAPK7 signaling. Biochem Bioph Res Co 474, 76–82 (2016).

81. Kulyté, A. et al. MicroRNA-27a/b-3p and PPARG regulate SCAMP3 through a feed-forward loop during adipogenesis. Sci Rep-uk 9, 13891 (2019).

82. Guo, Q., Chen, Y., Guo, L., Jiang, T. & Lin, Z. miR-23a/b regulates the balance between osteoblast and adipocyte differentiation in bone marrow mesenchymal stem cells. Bone Res 4, 16022 (2016).

83. Gu, C. et al. miR-27a attenuates adipogenesis and promotes osteogenesis in steroid-induced rat BMSCs by targeting PPARγ and GREM1. Sci Rep-uk 6, 38491 (2016).

84. Glass, C. K. & Olefsky, J. M. Inflammation and Lipid Signaling in the Etiology of Insulin Resistance. Cell Metab 15, 635–645 (2012).

85. Meikle, P. J. & Summers, S. A. Sphingolipids and phospholipids in insulin resistance and related metabolic disorders. Nat Rev Endocrinol 13, 79–91 (2017).

86. Mutt, S. J., Hyppönen, E., Saarnio, J., Järvelin, M.-R. & Herzig, K.-H. Vitamin D and adipose tissue—more than storage. Front Physiol 5, 228 (2014).

87. Raudvere, U. et al. g:Profiler: a web server for functional enrichment analysis and conversions of gene lists (2019 update). Nucleic Acids Res 47, W191–W198 (2019).

88. Yin, Y. et al. Impact of cytosine methylation on DNA binding specificities of human transcription factors. Science 356, eaaj2239 (2017).

89. Luo, C., Hajkova, P. & Ecker, J. R. Dynamic DNA methylation: In the right place at the right time. Science 361, 1336–1340 (2018).

90. Zhu, H., Wang, G. & Qian, J. Transcription factors as readers and effectors of DNA methylation. Nat Rev Genet 17, 551–565 (2016).

91. Heinz, S. et al. Simple Combinations of Lineage-Determining Transcription Factors Prime cis-Regulatory Elements Required for Macrophage and B Cell Identities. Mol Cell 38, 576–589 (2010).

92. Eckhardt, F. et al. DNA methylation profiling of human chromosomes 6, 20 and 22. Nat Genet 38, 1378–1385 (2006).

93. Wang, G. et al. MeDReaders: a database for transcription factors that bind to methylated DNA. Nucleic Acids Res 46, gkx1096-(2017).

94. White, U. A. & Stephens, J. M. Transcriptional factors that promote formation of white adipose tissue. Mol Cell Endocrinol 318, 10–14 (2010).

95. Suico, M. A., Shuto, T. & Kai, H. Roles and regulations of the ETS transcription factor ELF4/MEF. J Mol Cell Biol 1–10 (2016) doi:10.1093/jmcb/mjw051.

96. Pang, L., et al. miR-1275 inhibits adipogenesis via ELK1 and its expression decreases in obese subjects. J Mol Endocrinol 57, 33–43 (2016).

97. Wang, W. et al. Mediator MED23 Links Insulin Signaling to the Adipogenesis Transcription Cascade. Dev Cell 16, 764–771 (2009).

98. Li, S.-N. & Wu, J.-F. TGF-β SMAD signaling regulation of mesenchymal stem cells in adipocyte commitment. Stem Cell Res Ther 11, 41 (2020).

99. Sun, C. et al. Adipose Snail1 Regulates Lipolysis and Lipid Partitioning by Suppressing Adipose Triacylglycerol Lipase Expression. Cell Reports 17, 2015–2027 (2016).

100. Hou, X. et al. CDK6 inhibits white to beige fat transition by suppressing RUNX1. Nat Commun 9, 1023 (2018).

101. Pérez-Mancera, P. A. et al. Adipose tissue mass is modulated by SLUG (SNAI2). Hum Mol Genet 16, 2972–2986 (2007).

102. Leow, S. C. et al. The transcription factor SOX6 contributes to the developmental origins of obesity by promoting adipogenesis. Development 143, 950–961 (2016).

103. Chen, Q. et al. Fate decision of mesenchymal stem cells: adipocytes or osteoblasts? Cell Death Differ 23, 1128–1139 (2016).

104. Verdi, S. et al. TwinsUK: The UK Adult Twin Registry Update. Twin Res Hum Genet 22, 523–529 (2019).

105. Tsai, P.-C. et al. Smoking induces coordinated DNA methylation and gene expression changes in adipose tissue with consequences for metabolic health. Clin Epigenetics 10, 126 (2018).

106. Consortium, T. M. T. H. E. R. (MuTHER), et al. Mapping cis-and trans-regulatory effects across multiple tissues in twins. Nat Genet 44, 1084–1089 (2012).

107. Min, J. L. et al. Genomic and phenotypic insights from an atlas of genetic effects on DNA methylation. Nat Genet 53, 1311–1321 (2021).

108. Wheeler, E. et al. Impact of common genetic determinants of Hemoglobin A1c on type 2 diabetes risk and diagnosis in ancestrally diverse populations: A transethnic genome-wide meta-analysis. Plos Med 14, e1002383 (2017).

109. Hemani, G. et al. The MR-Base platform supports systematic causal inference across the human phenome. Elife 7, e34408 (2018).

110. Hemani, G., Tilling, K. & Smith, G. D. Orienting the causal relationship between imprecisely measured traits using GWAS summary data. Plos Genet 13, e1007081 (2017).

111. Chiang, S.-H. et al. Insulin-stimulated GLUT4 translocation requires the CAP-dependent activation of TC10. Nature 410, 944–948 (2001).

112. Chang, L., Adams, R. D. & Saltiel, A. R. The TC10-interacting protein CIP4/2 is required for insulin-stimulated Glut4 translocation in 3T3L1 adipocytes. Proc National Acad Sci 99, 12835–12840 (2002).

113. Wu, R. et al. A novel m6A reader Prrc2a controls oligodendroglial specification and myelination. Cell Res 29, 23–41 (2019).

114. Mendelson, M. M. et al. Association of Body Mass Index with DNA Methylation and Gene Expression in Blood Cells and Relations to Cardiometabolic Disease: A Mendelian Randomization Approach. Plos Med 14, e1002215 (2017).

115. Xu, X. et al. A genome-wide methylation study on obesity. Epigenetics 8, 522–533 (2013).

116. Feinberg, A. P. et al. Personalized Epigenomic Signatures That Are Stable Over Time and Covary with Body Mass Index. Sci Transl Med 2, 49ra67–49ra67 (2010).

117. Sayols-Baixeras, S. et al. DNA methylation and obesity traits: An epigenome-wide association study. The REGICOR study. Epigenetics 12, 909–916 (2017).

118. Demerath, E. W. et al. Epigenome-wide association study (EWAS) of BMI, BMI change and waist circumference in African American adults identifies multiple replicated loci. Hum Mol Genet 24, 4464–4479 (2015).

119. He, F. et al. Association between DNA methylation in obesity-related genes and body mass index percentile in adolescents. Sci Rep-uk 9, 2079 (2019).

120. Xu, K., Zhang, X., Wang, Z., Hu, Y. & Sinha, R. Epigenome-wide association analysis revealed that SOCS3 methylation influences the effect of cumulative stress on obesity. Biol Psychol 131, 63– 71 (2018).

121. Wang, X. et al. An epigenome-wide study of obesity in African American youth and young adults: novel findings, replication in neutrophils, and relationship with gene expression. Clin Epigenetics 10, 3 (2018).

122. Rönn, T., et al. Impact of age, BMI and HbA1c levels on the genome-wide DNA methylation and mRNA expression patterns in human adipose tissue and identification of epigenetic biomarkers in blood. Hum Mol Genet 24, 3792–3813 (2015).

123. Dick, K. J. et al. DNA methylation and body-mass index: a genome-wide analysis. Lancet 383, 1990–1998 (2014).

124. Benton, M. C. et al. An analysis of DNA methylation in human adipose tissue reveals differential modification of obesity genes before and after gastric bypass and weight loss. Genome Biol 16, 8 (2015).

125. Huang, R. C. et al. Genome-wide methylation analysis identifies differentially methylated CpG loci associated with severe obesity in childhood. Epigenetics 10, 995–1005 (2015).

126. Rzehak, P. et al. DNA-Methylation and Body Composition in Preschool Children: Epigenome-Wide-Analysis in the European Childhood Obesity Project (CHOP)-Study. Sci Rep-uk 7, 14349 (2017).

127. Fradin, D., et al. Genome-Wide Methylation Analysis Identifies Specific Epigenetic Marks In Severely Obese Children. Sci Rep-uk 7, 46311 (2017).

128. Dhana, K. et al. An Epigenome-Wide Association Study of Obesity-Related Traits. Am J Epidemiol 187, 1662–1669 (2018).

129. Aslibekyan, S. et al. Epigenome-wide study identifies novel methylation loci associated with body mass index and waist circumference. Obesity 23, 1493–1501 (2015).

130. Burgess, S. et al. Guidelines for performing Mendelian randomization investigations. Wellcome Open Res 4, 186 (2019).

131. Shema, E., Bernstein, B. E. & Buenrostro, J. D. Single-cell and single-molecule epigenomics to uncover genome regulation at unprecedented resolution. Nat Genet 51, 19–25 (2019).

132. Kelsey, G., Stegle, O. & Reik, W. Single-cell epigenomics: Recording the past and predicting the future. Science 358, 69–75 (2017).

133. Xu, X. et al. A CRISPR-based approach for targeted DNA demethylation. Cell Discov 2, 16009 (2016).

134. Vojta, A. et al. Repurposing the CRISPR-Cas9 system for targeted DNA methylation. Nucleic Acids Res 44, 5615–5628 (2016).

135. Liu, X. S. et al. Editing DNA Methylation in the Mammalian Genome. Cell 167, 233–247.e17 (2016).

136. Liu, X. S. et al. Rescue of Fragile X Syndrome Neurons by DNA Methylation Editing of the FMR1 Gene. Cell 172, 979–992.e6 (2018).

137. Carswell, K. A., Lee, M.-J. & Fried, S. K. Human Cell Culture Protocols. Methods Mol Biology 806, 203–214 (2011).

138. Spalding, K. L. et al. Dynamics of fat cell turnover in humans. Nature 453, 783–787 (2008).

139. Aryee, M. J. et al. Minfi: a flexible and comprehensive Bioconductor package for the analysis of Infinium DNA methylation microarrays. Bioinformatics 30, 1363–1369 (2014).

140. Lehne, B. et al. A coherent approach for analysis of the Illumina HumanMethylation450 BeadChip improves data quality and performance in epigenome-wide association studies. Genome Biol 16, 37 (2015).

141. Leek, J. T., Johnson, W. E., Parker, H. S., Jaffe, A. E. & Storey, J. D. The sva package for removing batch effects and other unwanted variation in high-throughput experiments. Bioinformatics 28, 882–883 (2012).

142. Love, M. I., Huber, W. & Anders, S. Moderated estimation of fold change and dispersion for RNA-seq data with DESeq2. Genome Biol 15, 550 (2014).

143. Chen, Y. et al. Discovery of cross-reactive probes and polymorphic CpGs in the Illumina Infinium HumanMethylation450 microarray. Epigenetics 8, 203–209 (2013).

144. Price, E. M. et al. Additional annotation enhances potential for biologically-relevant analysis of the Illumina Infinium HumanMethylation450 BeadChip array. Epigenet Chromatin 6, 4 (2013).

145. Lehne, B. et al. A coherent approach for analysis of the Illumina HumanMethylation450 BeadChip improves data quality and performance in epigenome-wide association studies. Genome Biol 16, 37 (2015).

146. Tahir, R. A., Zheng, D., Nazir, A. & Qing, H. A review of computational algorithms for CpG islands detection. J Biosciences 44, 143 (2019).

147. Delaneau, O., Marchini, J. & Zagury, J.-F. A linear complexity phasing method for thousands of genomes. Nat Methods 9, 179–181 (2012).

148. Marchini, J., Howie, B., Myers, S., McVean, G. & Donnelly, P. A new multipoint method for genome-wide association studies by imputation of genotypes. Nat Genet 39, 906–913 (2007).

149. Brägelmann, J. & Bermejo, J. L. A comparative analysis of cell-type adjustment methods for epigenome-wide association studies based on simulated and real data sets. Brief Bioinform 20, 2055– 2065 (2018).

150. McGregor, K. et al. An evaluation of methods correcting for cell-type heterogeneity in DNA methylation studies. Genome Biol 17, 84 (2016).

151. Kaushal, A. et al. Comparison of different cell type correction methods for genome-scale epigenetics studies. Bmc Bioinformatics 18, 216 (2017).

152. Yu, G., Wang, L.-G. & He, Q.-Y. ChIPseeker: an R/Bioconductor package for ChIP peak annotation, comparison and visualization. Bioinformatics 31, 2382–2383 (2015).

153. Subramanian, A. et al. Gene set enrichment analysis: A knowledge-based approach for interpreting genome-wide expression profiles. P Natl Acad Sci Usa 102, 15545–15550 (2005).

154. Liberzon, A. et al. Molecular signatures database (MSigDB) 3.0. Bioinformatics 27, 1739–1740 (2011).

155. Isidor, M. S. et al. An siRNA-based method for efficient silencing of gene expression in mature brown adipocytes. Adipocyte 5, 175–185 (2015).

156. Anunciado-Koza, R. P. et al. Diet-induced adipose tissue expansion is mitigated in mice with a targeted inactivation of mesoderm specific transcript (Mest). Plos One 12, e0179879 (2017).

157. Morigny, P., Boucher, J., Arner, P. & Langin, D. Lipid and glucose metabolism in white adipocytes: pathways, dysfunction and therapeutics. Nat Rev Endocrinol 17, 276–295 (2021).

