## Supplementary Figures for "Integrative genomic analyses in adipocytes implicate DNA methylation in human obesity and diabetes"

**Supplementary Figure 1**
**Study Design.**

**Supplementary Figure 2A**
**Effect of genetic factors on methylation-phenotype associations in subcutaneous adipocytes.**

**Supplementary Figure 2B**
**Effect of genetic factors on methylation-phenotype associations in visceral adipocytes.**

**Supplementary Figure 3A**
**Evaluation of potential adipocyte impurity by contaminating gene expression in subcutaneous adipocytes.**

**Supplementary Figure 3B**
**Evaluation of potential adipocyte impurity by contaminating gene expression in visceral adipocytes.**

**Supplementary Figure 4**
**Evaluation of potential adipocyte impurity by SVA.**

**Supplementary Figure 5**
**Genomic annotation of subcutaneous and visceral sentinels.**

**Supplementary Figure 6**
**DNA methylation-target gene associations in human subcutaneous and visceral adipocytes annotated by Roadmap chromatin states.**

**Supplementary Figure 7**
**Nodal plots of enriched pathways and geneset of target genes associated with DNA methylation sentinels at FDR<0.01 in subcutaneous and visceral adipocytes.**

**Supplementary Figure 8**
**Genomic and functional annotation of enriched TF motifs in subcutaneous adipocytes.**

**Supplementary Figure 9**
**Adipocyte functional studies replication.**

**Supplementary Figure 10**
**Human adipocyte RNA sequencing quality control analyses.**

**Supplementary Table 1**
**Participant characteristics**

**Supplementary Table 2**
**Subcutaneous DNA methylation sentinels associated with extreme human obesity in discovery and replication analyses.**

**Supplementary Table 3**
**Visceral DNA methylation sentinels associated with extreme human obesity in discovery and replication analyses.**

**Supplementary Table 4**
**Cross-depot comparisons of significant DNA methylation sentinels associated with extreme obesity.**

**Supplementary Table 5**

**Positional overlap of DNA methylation sentinels with human adiposity and metabolic disease GWAS SNPs.**

**Supplementary Table 6**
**Associations between genic sentinels and flanking/overlapping cis- target genes in subcutaneous adipocytes.**

**Supplementary Table 7**
**Associations between genic sentinels and flanking/overlapping cis- target genes in visceral adipocytes.**

**Supplementary Table 8**
**Associations between non-genic sentinels and functionally assigned cis- target genes in subcutaneous adipocytes.**

**Supplementary Table 9**
**Associations between non-genic sentinels and functionally assigned cis- target genes in visceral adipocytes.**

**Supplementary Table 10**
**Associations between non-genic sentinels and TAD assigned cis- target genes in subcutaneous adipocytes.**

**Supplementary Table 11**
**Associations between non-genic sentinels and TAD assigned cis- target genes in visceral adipocytes.**

**Supplementary Table 12**
**Pathway and gene set analyses of the nearest cis- genes to each subcutaneous and visceral DNA methylation sentinel.**

**Supplementary Table 13**
**Pathway and gene set analyses of cis- target genes associated with subcutaneous and visceral methylation sentinels.**

**Supplementary Table 14**
**Enriched transcription factor binding motifs within +/-150-bp of the subcutaneous DNA methylation sentinels.**

**Supplementary Table 15**
**Transcription factors predicted to bind at enriched transcription factor binding motifs in subcutaneous adipocytes.**

**Supplementary Table 16**
**Putative transcription factor binding motifs within +/-150-bp of the visceral DNA methylation sentinels.**

**Supplementary Table 17**
**Mendelian Randomization analyses investigating the causal effects of adipose tissue DNA methylation on human obesity, obesity-induced metabolic disturbances and T2D.**

**Supplementary Table 18**
**Replication of human adipose tissue mQTLs with MR evidence of disease causation in human subcutaneous and visceral adipocytes.**

**Supplementary Table 19**
**Primers used in RT-qPCR studies of gene expression in adipocytes.**

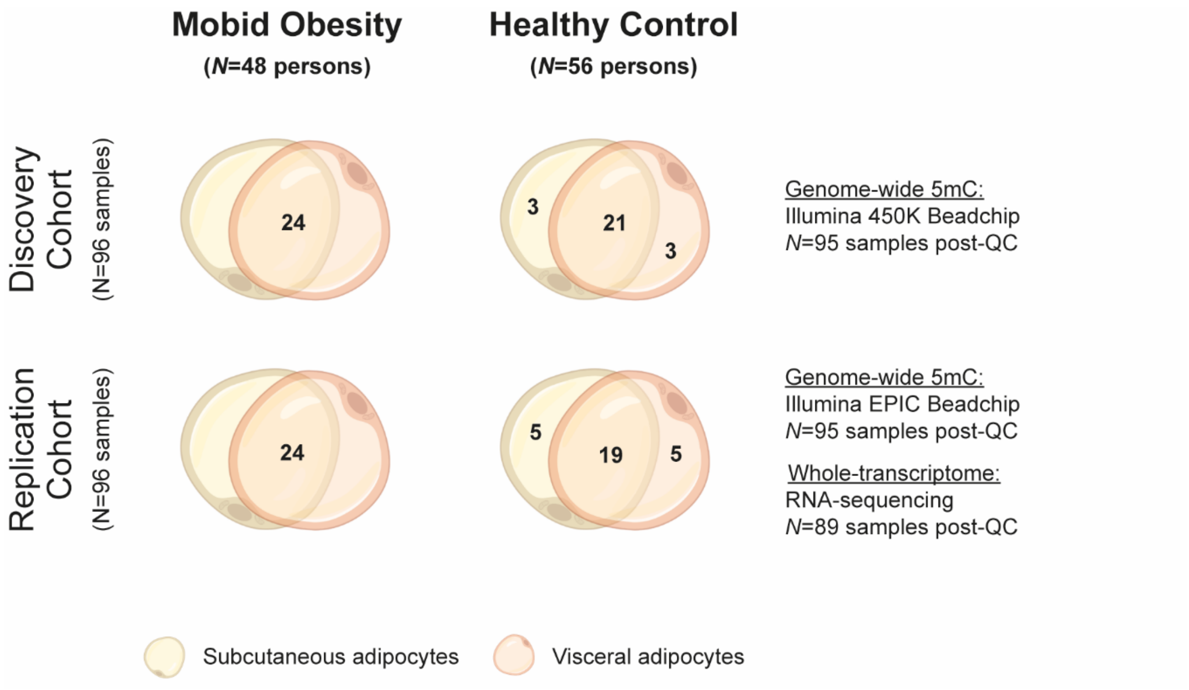


**Supplementary Figure 1: Study Design.**

We evaluated 192 adipocyte samples, half subcutaneous and half visceral, from people with extreme obesity and healthy controls in separate discovery and replication cohorts. In the discovery cohort, 24 obese people and 21 controls provided paired subcutaneous and visceral adipocyte samples. A further 3 controls provided just subcutaneous and 3 controls provided just visceral adipocytes. Similarly, in the replication cohort, 24 obese people and 19 controls provided paired subcutaneous and visceral adipocyte samples, with 10 further controls providing 5 subcutaneous and 5 visceral samples only. Genome-wide DNA methylation was characterised using the Illumina Methylation450 Beadchip in the 96 discovery samples, and the Illumina EPIC Beadchip in the 96 replication samples. Two methylation samples that failed quality control analyses were removed, one in each of the discovery and replication cohorts. RNA sequencing was carried out in the 96 replication samples, and 7 samples that failed sequencing or quality control analyses were removed.

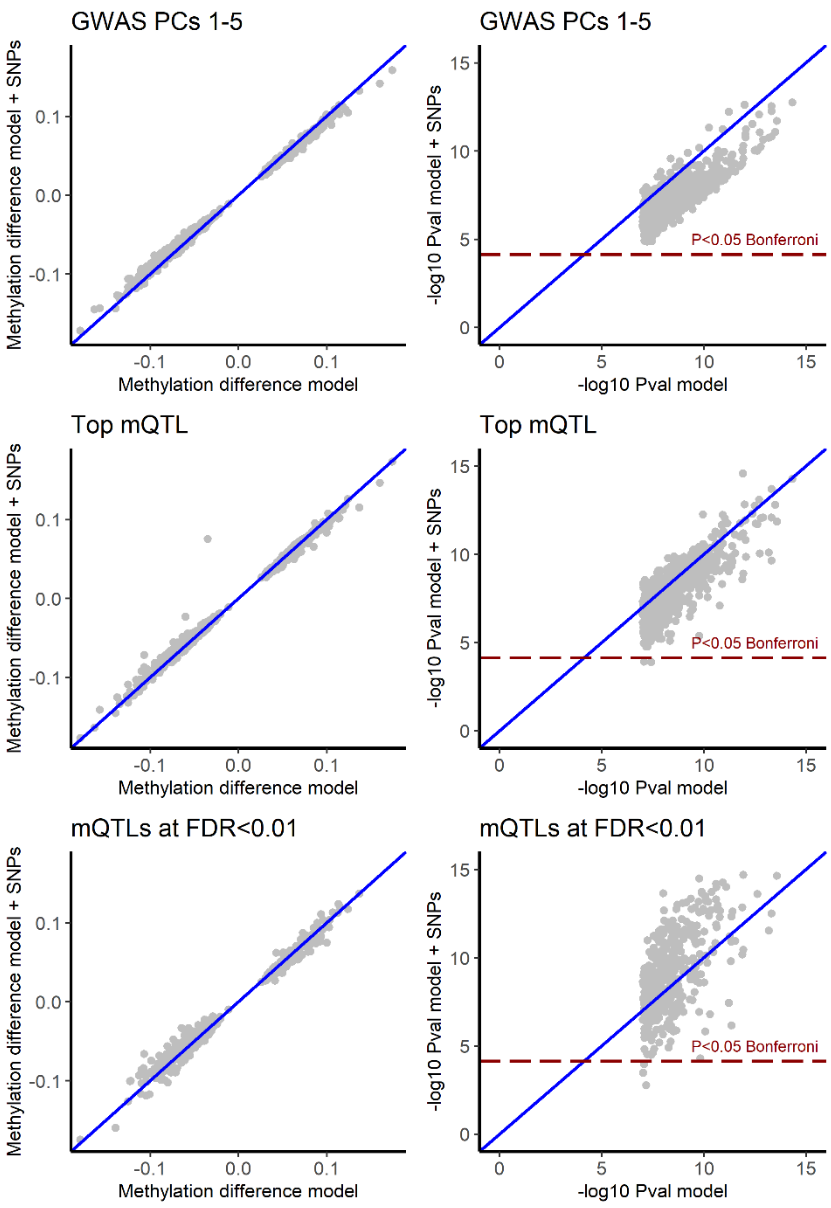


**Supplementary Figure 2A: Effect of genetic factors on methylation-phenotype associations in subcutaneous adipocytes.**

Associations between subcutaneous adipocyte DNA methylation and extreme obesity at each sentinel site were compared in: i. the base model (X-axis: methylation difference model); and ii. the base adjusted for the effects of genetic variants (Y-axis: methylation difference model + SNPs, combined discovery and replication cohorts). Genetic factors did not systematically affect the relationships between DNA methylation and obesity, indicating that the majority of methylation-phenotype associations are likely to be environmentally driven. Top panels: adjustment for the first 5 principal components from PCA analysis of >8M genome-wide SNPs carried out in all study participants. Middle panels: adjustment for the genotype dose of the top cis-SNP (±500-kb) associated with change in methylation at each sentinel site (Top mQTL, additive model). Bottom panels: adjustment for the genotype dose of all cis-SNPs (±500-kb) associated with change in sentinel methylation at FDR<0.01 (additive model). Left panels: association effect sizes (betas). Right panels: -log10 pvalues for association. Blues lines: Null of no systematic difference. Dark red lines: Significance threshold (P<0.05, Bonferroni adjusted for the number of sentinels).

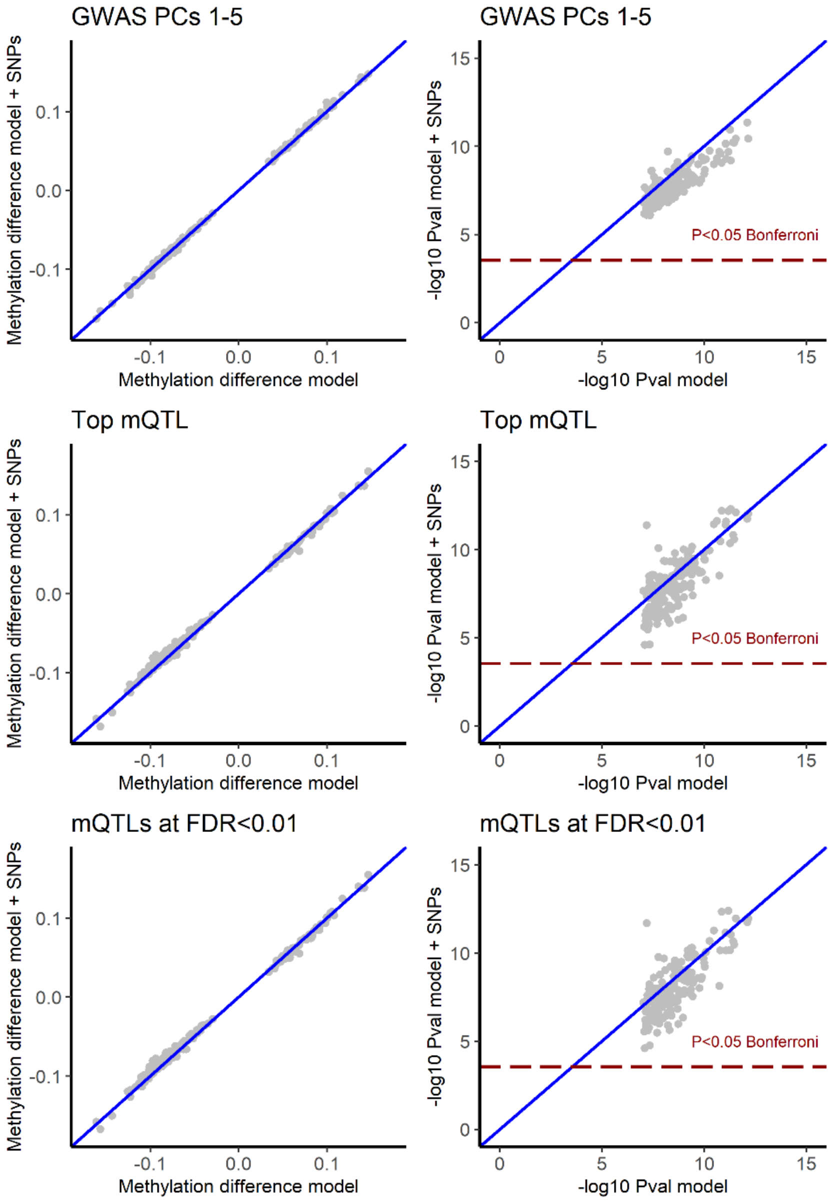


**Supplementary Figure 2B: Effect of genetic factors on methylation-phenotype associations in visceral adipocytes.**

Comparisons of associations between visceral adipocyte DNA methylation and extreme obesity at each sentinel site in: i. the base model (X-axis: methylation difference model); and ii. the base adjusted for the effects of genetic variants (Y-axis: methylation difference model + SNPs, combined discovery and replication cohorts). As in subcutaneous adipocytes, genetic factors did not systematically affect DNA methylation-obesity associations, indicating underlying environmental origins. Top panels: adjusted for the first 5 principal components from PCA analysis of >8M SNPs carried out in all study participants. Middle panels: adjusted for the genotype dose of the top cis-SNP (±500-kb) associated with change in methylation at each sentinel site (Top mQTL, additive model). Bottom panels: adjusted for the genotype dose of all cis-SNPs (±500-kb) associated with change in sentinel methylation at FDR<0.01 (additive model). Left panels: association effect sizes (betas). Right panels: -log10 pvalues for association. Blues lines: Null of no systematic difference. Dark red lines: Significance threshold (P<0.05, Bonferroni adjusted for the number of sentinels).

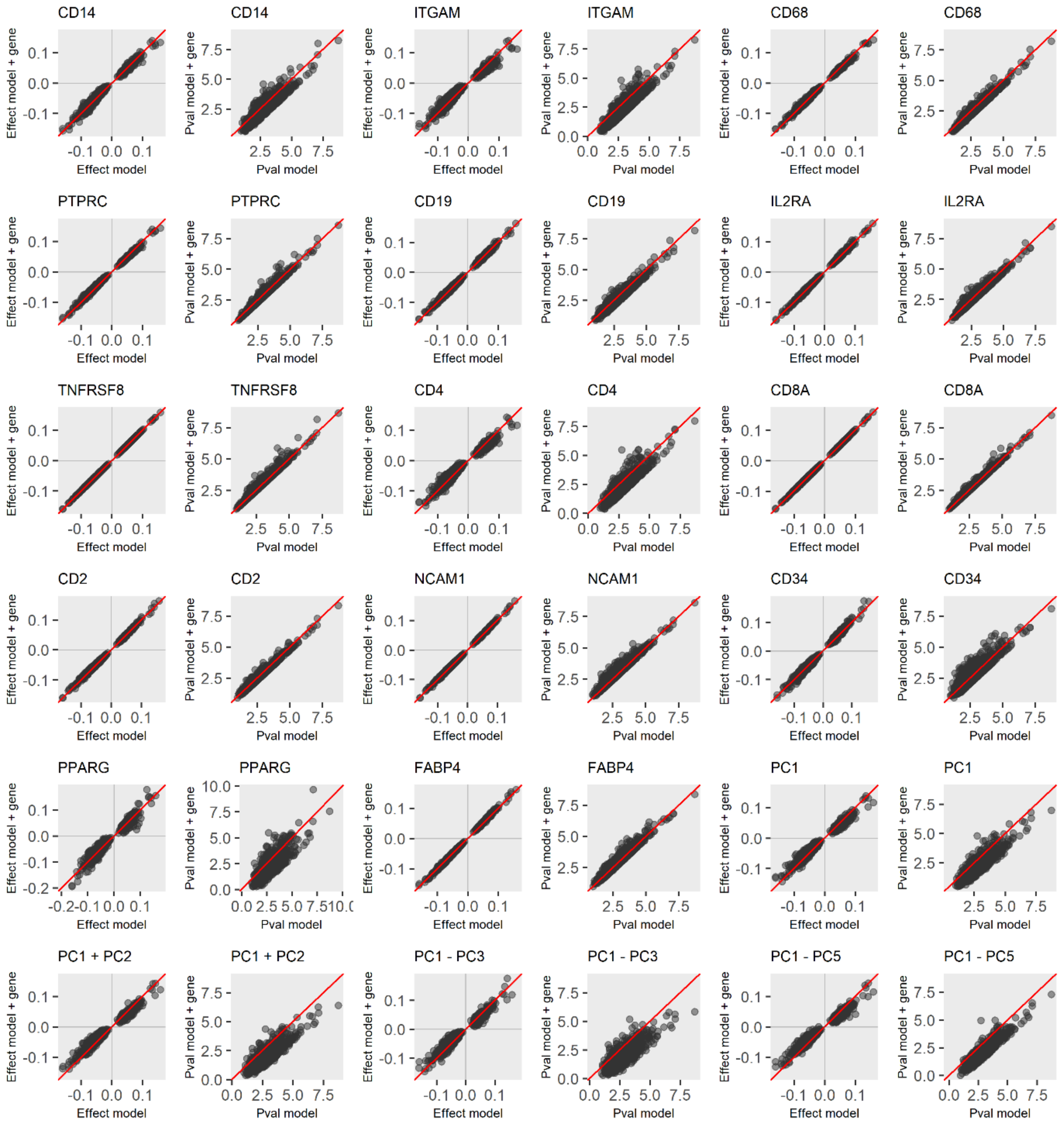


**Supplementary Figure 3A: Evaluation of potential adipocyte impurity by contaminating gene expression in subcutaneous adipocytes.**

Gene expression results from RNA sequencing were used to evaluate the effects of potential contaminating cells (impurity) on DNA methylation-obesity associations by comparing base models without and with adjustment for gene expression levels (subcutaneous, replication cohort). Effects of 14 individual genes were evaluated (12 potential contaminating and 2 control genes), as were principal components (PCs) from PCA analyses of all 12 potential contaminating cell genes. No systematic effects on DNA methylation-obesity associations at the 691 subcutaneous sentinel sites were observed. Red lines: Null of no systematic difference. Identifiers: *CD14* (monocyte/macrophage); *ITGAM* (CD11b, broad immune cell); CD68 (monocyte/macrophage); *PTPRC* (CD45, broad immune cell); *CD19* (B lymphocyte); *IL2RA* (T lymphocyte); *TNFRSF8* (CD39, broad lymphocyte); *CD4* (T lymphocyte subtype); *CD8A* (T lymphocyte subtype); *CD2* (T lymphocyte and NK cell); *NCAM1* (NK cell); *CD34* (endothelial and precursor cell); *PPARG* (adipocyte); *FABP4* (adipocyte); PC1-PC5 (principal components from PCA analysis of the 12 immune and stromovascular genes). Analyses were carried out with variance stabilizing gene expression counts.

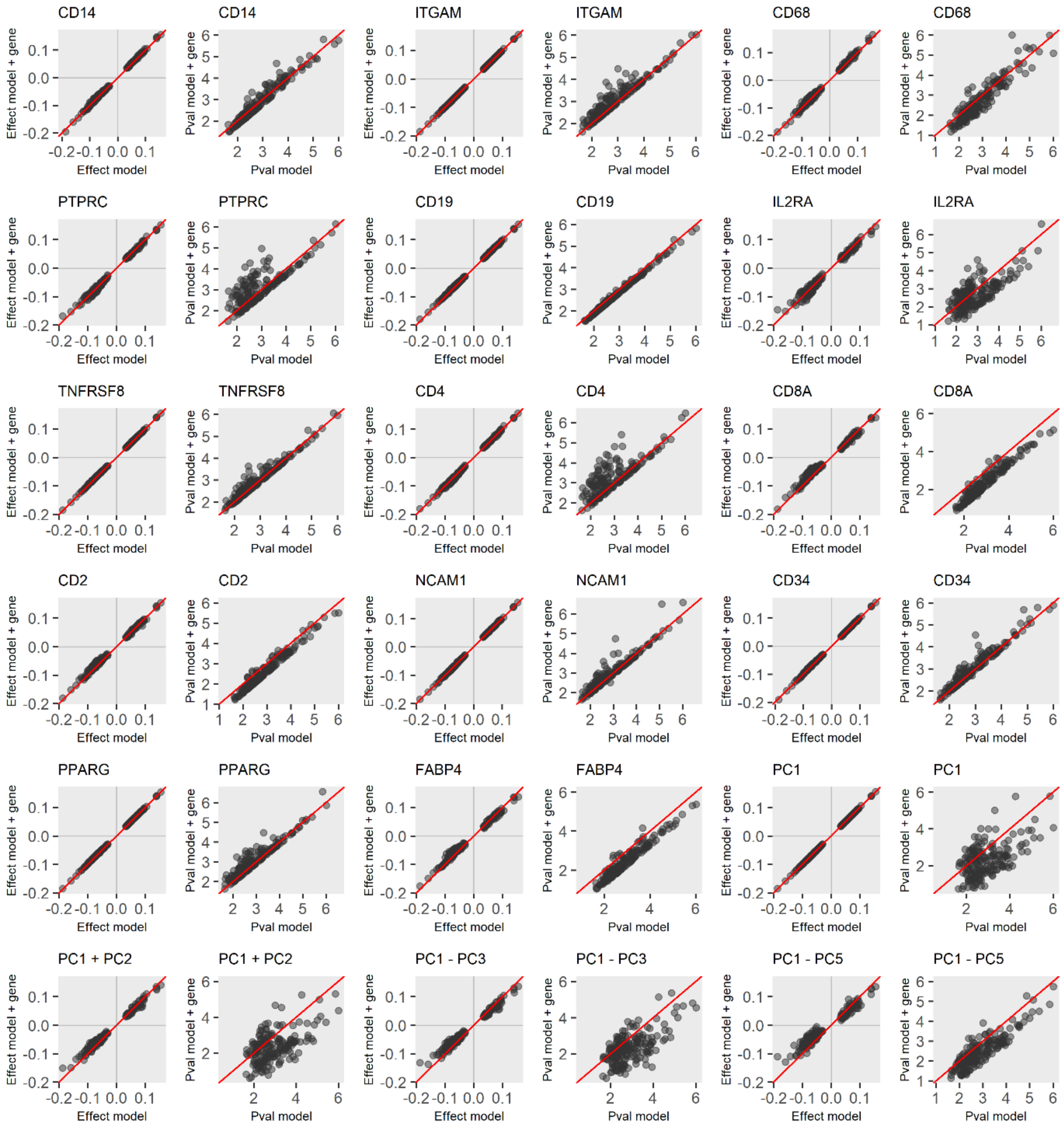


**Supplementary Figure 3B: Evaluation of potential adipocyte impurity by contaminating gene expression in visceral adipocytes.**

Effect of contaminating cell gene expression results from RNA sequencing (impurity) on DNA methylation-obesity associations (visceral replication cohort, 12 potential contaminating and 2 control genes, and principal components (PCs) from PCA analyses of all 12 potential contaminating cell genes). No systematic effects were observed at the 173 visceral sentinel sites. Red lines: Null of no systematic difference. Identifiers: *CD14* (monocyte/macrophage); *ITGAM* (CD11b, broad immune cell); CD68 (monocyte/macrophage); *PTPRC* (CD45, broad immune cell); *CD19* (B lymphocyte); *IL2RA* (T lymphocyte); *TNFRSF8* (CD39, broad lymphocyte); *CD4* (T lymphocyte subtype); *CD8A* (T lymphocyte subtype); *CD2* (T lymphocyte and NK cell); *NCAM1* (NK cell); *CD34* (endothelial and precursor cells); *PPARG* (adipocyte); *FABP4* (adipocyte); PC1-PC5 (principal components from PCA analysis of the 12 immune and stromovascular genes). Analyses carried out with variance stabilizing gene expression counts.

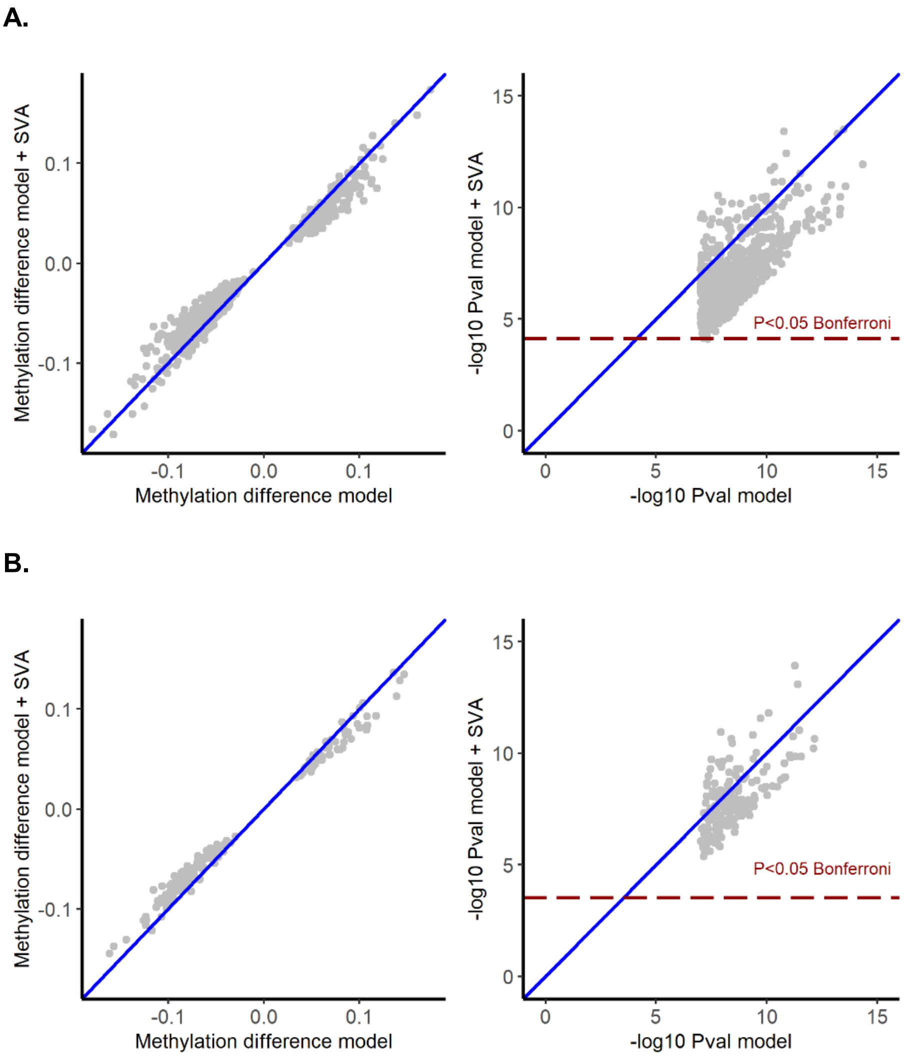


**Supplementary Figure 4: Evaluation of potential adipocyte impurity by SVA.**

Surrogate variable analysis was used to evaluate the effects of unknown confounders, in particular potential contaminating cell impurity, on associations between DNA methylation and extreme obesity in **A.** subcutaneous and **B.** visceral adipocytes (combined discovery and replication cohorts). Inclusion of SV1 and SV2 in the base models did not systematically alter methylation-phenotype effect sizes, and the majority of sentinels remained significantly associated with obesity after adjustment for SV1 and SV2 in the base model. Left panels: association betas. Right panels: -log10 pvalues. Blues lines: Null of no systematic difference. Dark red lines: Significance threshold (P<0.05, Bonferroni adjusted for the number of sentinels).

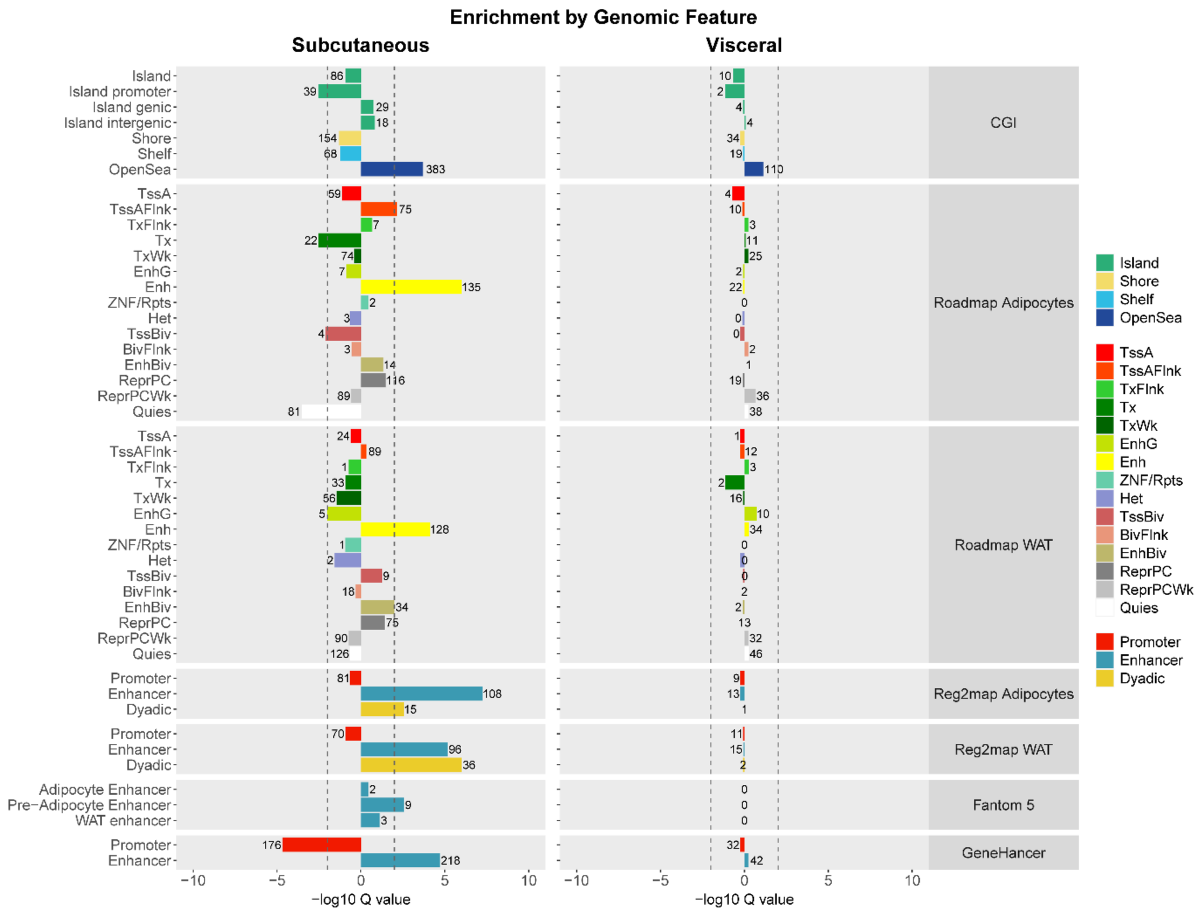


**Supplementary Figure 5: Genomic annotation of subcutaneous and visceral sentinels.**

Localisation of DNA methylation sentinels in subcutaneous and visceral adipocytes to functional/active genomic regions. Subcutaneous sentinels (N=691) were enriched in enhancers from various datasets and in CpG sparse genomic regions (open sea) but were underrepresented in promoter CGIs. Visceral sentinels (N=173) showed generally similar trends but did not reach statistical significance in any feature. CGI: CpG island track from UCSC. Presented as -log10 enrichment or depletion Q value, and number of observed counts, for each feature. Roadmap adipocytes and WAT: human adipocyte (E025) and adipose tissue nuclei chromatin states from the Roadmap epigenomics consortium (E063). Reg2map adipocytes and WAT: human adipocyte and adipose tissue regulatory features called using Roadmap and Encode epigenomes (-log10 pvalue ≥10). Fantom 5: human enhancer tracks called from Fantom 5 CAGE enhancer-promoter co-expression (ref). GeneHancer: multifaceted human enhancer-target gene inference database.

TssA: Active TSS. TssAFlnk: Flanking Active TSS. TxFlnk: Transcr. at gene 5' and 3'. Tx: Strong transcription. TxWk: Weak transcription. EnhG: Genic enhancers. Enh: Enhancers. ZNF/Rpts: ZNF genes & repeats. Het: Heterochromatin. TssBiv: Bivalent/Poised TSS. BivFlnk: Flanking Bivalent TSS/Enh. EnhBiv: Bivalent Enhancer. ReprPC: Repressed PolyComb. ReprPCWk: Weak Repressed PolyComb. Quies: Quiescent/Low. Promoter: inferred promoter. Enhancer: inferred enhancer. Dyadic: inferred promoter/enhancer.

**A.**

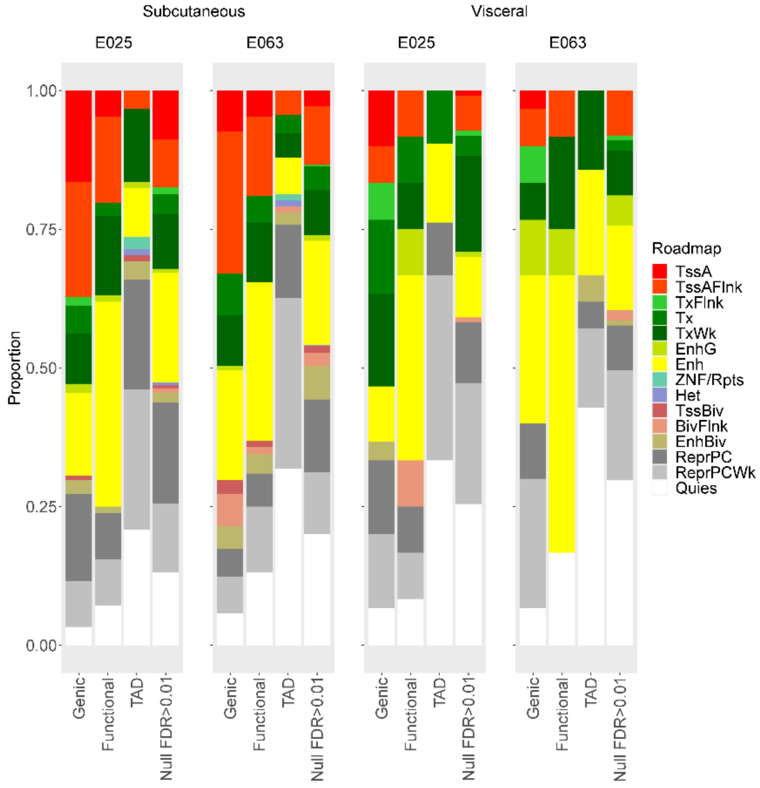


**B.**

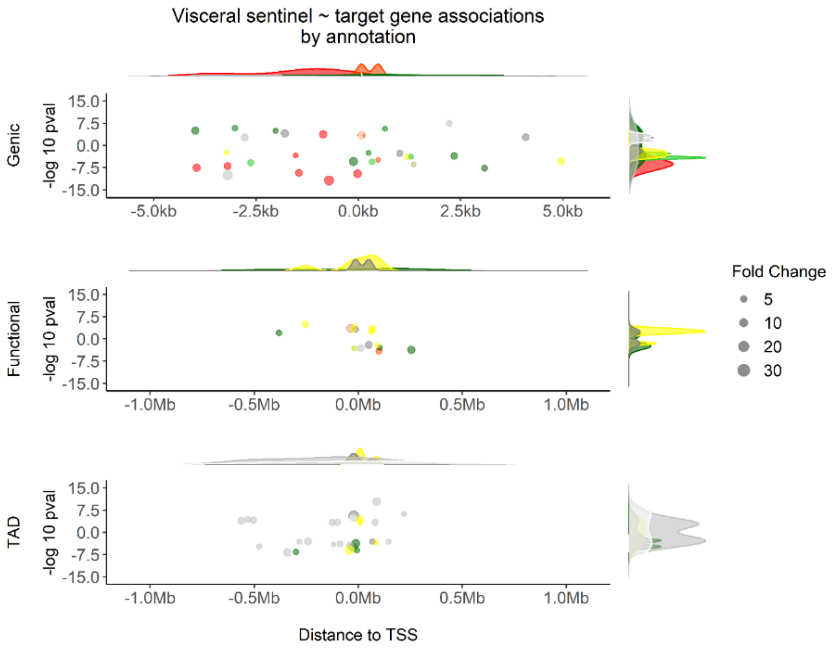


**Supplementary Figure 6: DNA methylation-target gene associations in human subcutaneous and visceral adipocytes annotated by Roadmap chromatin states.**

**A.** Subcutaneous and visceral adipocyte sentinel-target gene associations at FDR<0.01 grouped by annotation method and presented as proportion of associations in each Roadmap chromatin state. Genic: sentinel in a promoter, 5/3'UTR or exon. Functional: intronic/intergenic sentinel sharing a functional interaction with a distal target gene. TAD: intronic/intergenic sentinels and distal target genes sharing a topologically associated domain in human adipocytes. Null: methylation-target gene associations at FDR>0.01. E025: Human adipocytes. E063: human adipose tissue nuclei. All sentinel DNA methylation-target gene expression analyses were carried out in combined subcutaneous and visceral adipocyte samples.

TssA: Active TSS. TssAFlnk: Flanking Active TSS. TxFlnk: Transcr. at gene 5' and 3'. Tx: Strong transcription. TxWk: Weak transcription. EnhG: Genic enhancers. Enh: Enhancers. ZNF/Rpts: ZNF genes & repeats. Het: Heterochromatin. TssBiv: Bivalent/Poised TSS. BivFlnk: Flanking Bivalent TSS/Enh. EnhBiv: Bivalent Enhancer. ReprPC: Repressed PolyComb. ReprPCWk: Weak Repressed PolyComb. Quies: Quiescent/Low.

**B.** Visceral adipocyte sentinel-target gene associations at FDR<0.01, grouped by annotation method, presented as distance between the sentinel and its target gene TSS, and coloured by roadmap chromatin state (E025 adipocytes). Scatter plots of distance and -log 10 pvalue for association split according to direction of effect (methylation-expression). Density plots of each sentinel-target gene association relative to: i. distance (top); and ii. -log10 pvalue (right border). Fold change: expression fold change per unit change in methylation.

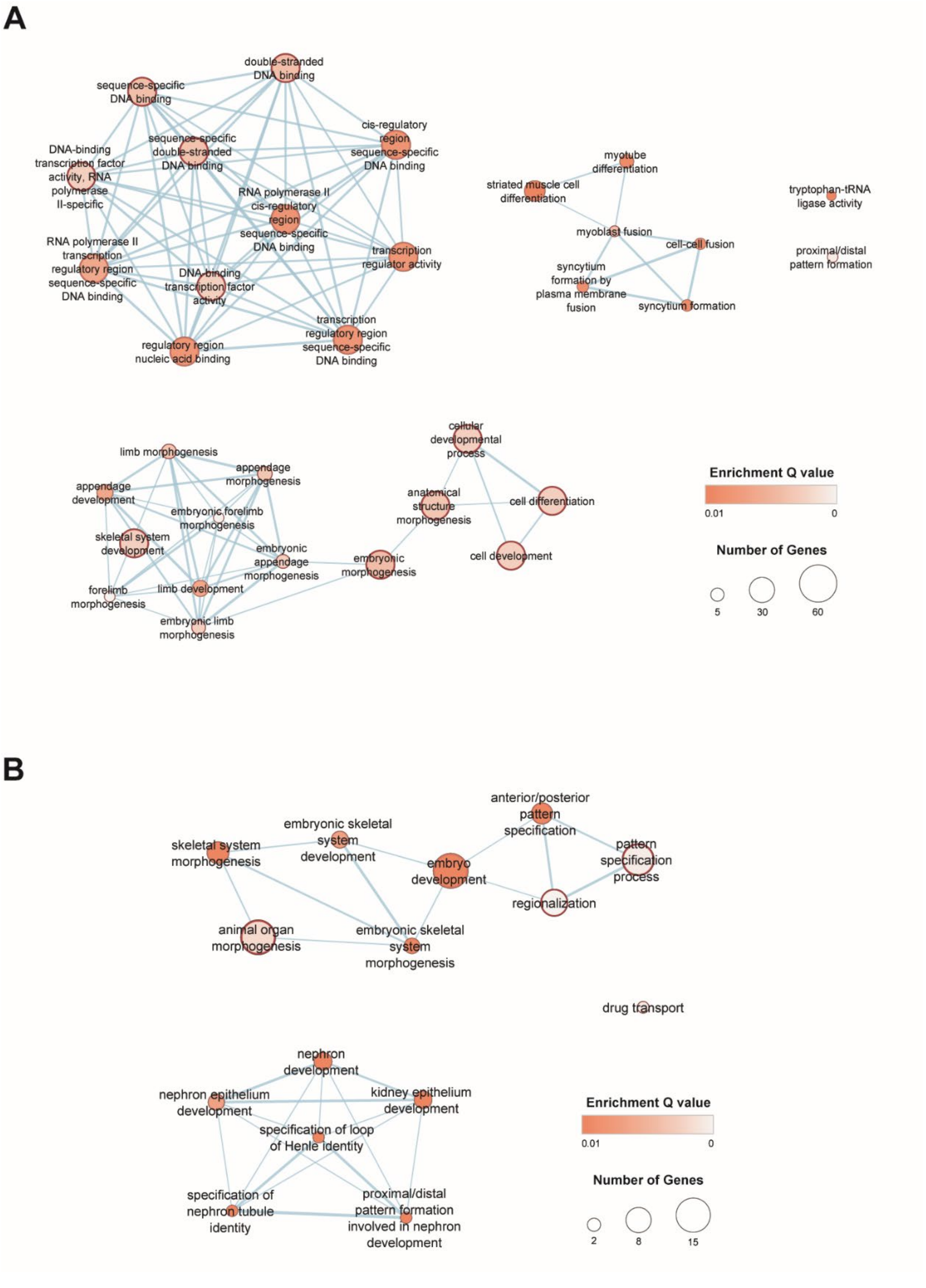


**Supplementary Figure 7: Nodal plots of enriched pathways and geneset of target genes associated with DNA methylation sentinels at FDR<0.01 in subcutaneous and visceral adipocytes (gProfiler, Enrichment Map).**

**A.** Subcutaneous: 33 enriched pathways/genesets clustered into transcriptional control and tissue development genesets, and cell, muscle and limb/morphogenesis subclusters. **B.** Visceral: 15 enriched pathways/genesets clustered into embryonic development/body patterning and nephronic development genesets. Coloured by enrichment Q value and sized by number of geneset members.

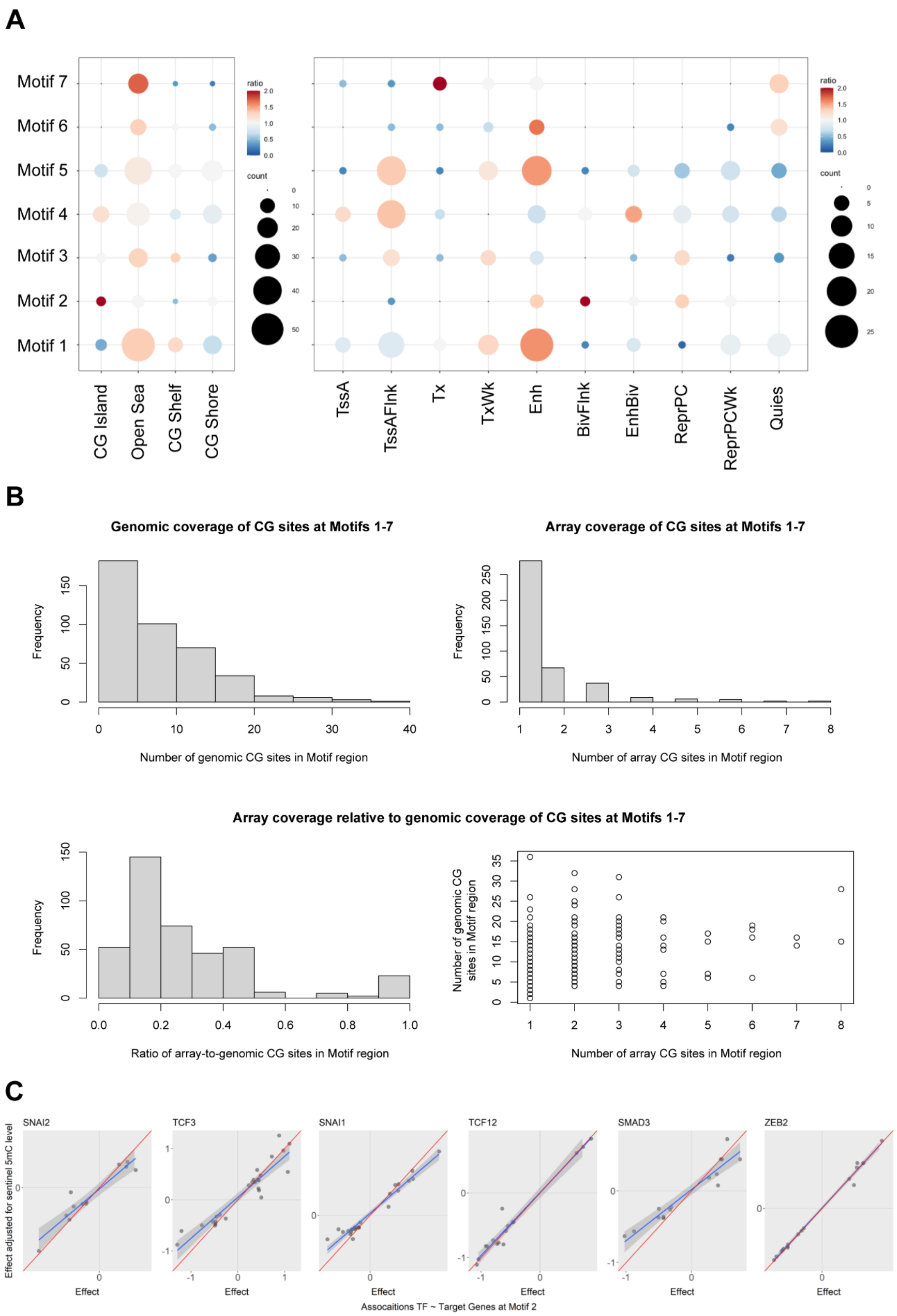


**Supplementary Figure 8: Genomic and functional annotation of enriched TF motifs in subcutaneous adipocytes.**

**A.** Genomic annotation of TF Motifs 1-7 in human CGIs and human adipocyte Roadmap chromatin states. Sized by the observed number of sentinel-motif pairs (count), and coloured by the ratio of observed relative to the expected counts (permuted background). 5 of 15 Roadmap states with low observed counts in all 7 motifs are not presented.

TssA: Active TSS. TssAFlnk: Flanking Active TSS. Tx: Strong transcription. TxWk: Weak transcription. Enh: Enhancers. BivFlnk: Flanking Bivalent TSS/Enh. EnhBiv: Bivalent Enhancer. ReprPC: Repressed PolyComb. ReprPCWk: Weak Repressed PolyComb. Quies: Quiescent/Low.

**B.** Density of genomic CG sites at Motifs 1 to 7, and coverage of these CG sites on the Illumina HumanMethylation450 array. Top left panel: Frequency histogram of the number of genomic CG sites present within the +/-150-bp regions flanking Motifs 1-7. Top right panel: Frequency histogram of the number of CG sites present on the Illumina Humanmethylation450 array within the +/-150-bp regions flanking Motifs 1-7. Bottom panels: Coverage of genomic CG sites within the +/-150-bp regions flanking Motifs 1-7 by the Illumina Humanmethylation450 array, presented as the ratios (left) and counts (right).

**C.** Association between expression of 6 TFs predicted to bind to Motif 2, and expression of the predicted target genes of each sentinel methylation site corresponding to Motif 2 (combined subcutaneous and visceral adipocyte samples). X-axis: association betas without adjustment for sentinel methylation level. Y-axis: association betas with adjustment for sentinel methylation level. Adjustment for sentinel methylation level systematically impacted TF-target gene association relationships involving *SNAI2*, *TCF3*, *SNAI1* and *SMAD3*, but not *TCF12* and *ZEB2*.

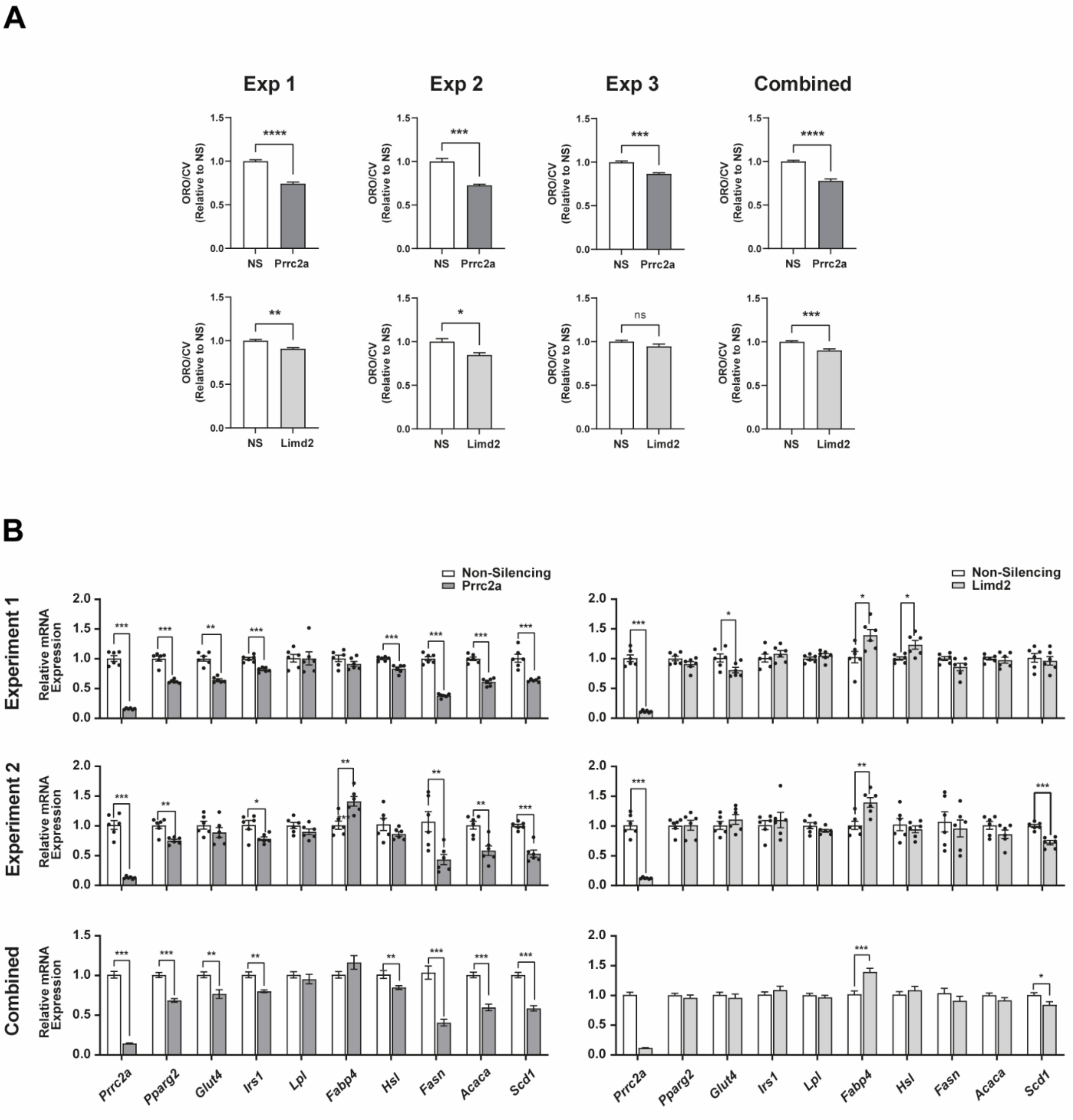


**Supplementary Figure 9: Adipocyte functional studies replication.**

Studies of lipid accumulation and expression of key genes involved in adipocyte metabolism at day 6 of differentiation, in 3T3-L1 adipocytes transfected with siRNA against *Prrc2a*, *Limd2* or non-silencing (NS) control at day 2 after initiation of differentiation.

**A.** Effects of *Prrc2a* and *Limd2* knockdown on lipid accumulation, assessed using spectrophotometry of eluted ORO stain normalised for cell number (crystal violet, CV, N=4 per condition). Results are shown for three replicate experiments (Exp. 1-3) and for the combined replicates.

**B.** Effects of *Prrc2a* and *Limd2* knockdown on the expression of genes involved in adipocyte differentiation (*Pparg*), insulin signalling (*Glut4*, *Irs1*), lipid uptake (*Lpl*), lipid storage (*Fasn*, *Acaca*, *Scd1*) and lipid mobilisation (*Fabp4*, *Hsl*, N=6 per condition). Real-time qPCR values were normalised to housekeeping genes (*Nono*, *Ywhaz*). Results are shown for two replicate experiments (Experiment 1-2, with individual data points) and for the combined replicates.

All values are presents as mean ± SEM relative to NS control. *P<0.05, **P<0.01, ***P<0.001, ****P<0.0001 versus NS control (Student's t-test).

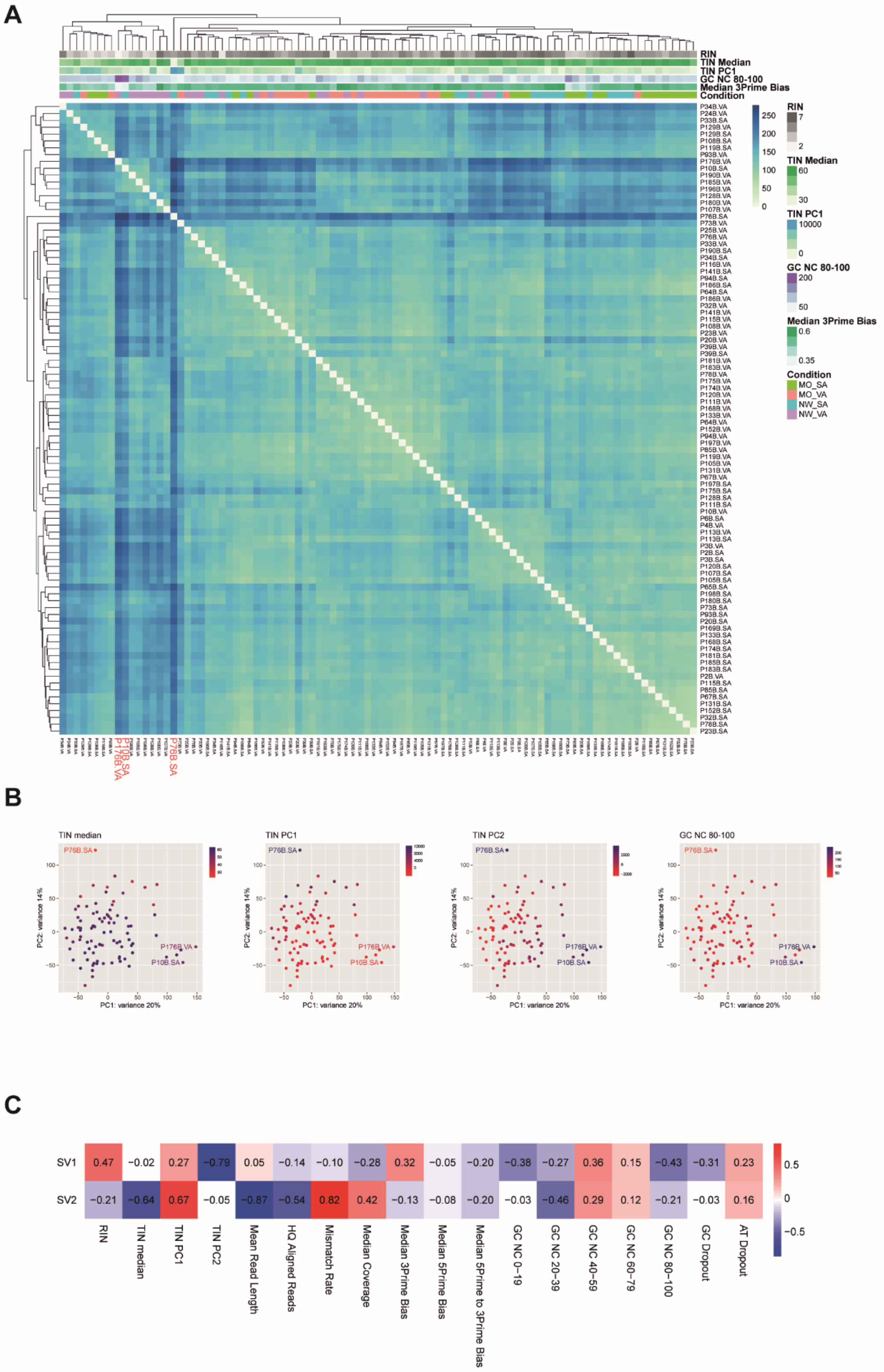


**Supplementary Figure 10: Human adipocyte RNA sequencing quality control analyses.**

**A.** Heatmap of between sample distance matrix (euclidean) of whole-transcriptome results (variance stabilising transformed counts, 92 subcutaneous and visceral adipocyte samples with RNA sequencing results at >15M assigned reads). Coloured by RNA integrity (RIN), transcript integrity (TIN Median and PC1), GC bias (GC NC 80-100), coverage bias (Median 3Prime Bias) and sample type. MO_SA: obese subcutaneous. NW_SA: control subcutaneous. MO_VA: obese visceral. NW_VA: control visceral.

**B.** Principal components 1 and 2 from principal component analysis of whole-transcriptome results (variance stabilising transformed counts, 92 subcutaneous and visceral adipocyte samples). Coloured by sequencing quality control metric: TIN (Median, PC1 and PC2) and GC bias (GC NC 80-100).

**C.** Surrogate variable analysis of whole-transcriptome results (variance stabilising transformed counts, 92 subcutaneous and visceral adipocyte samples). Pairwise correlation of resulting surrogate variables 1 and 2 with measures of sample quality and sequencing quality (Pearsons): RNA quality (RIN); read and transcript length and quality (TIN Median, PC1 and PC2, Mean Read Length, HQ (high-quality) Aligned Reads, Mismatch Rate); sequencing coverage (Median Coverage, 3Prime, 5Pime, 5Prime to 3Prime Bias) and sequencing GC bias (GC NC 0-100, GC Dropout, AT Dropout).
